# Blunted anticipation but not consummation of food rewards in depression

**DOI:** 10.1101/2024.03.26.24304849

**Authors:** Corinna Schulz, Johannes Klaus, Franziska Peglow, Sabine Ellinger, Anne Kühnel, Martin Walter, Nils B. Kroemer

**Author notes:** **Corresponding author** Prof. Dr. Nils B. Kroemer, Venusberg Campus 1, 53127 Bonn, Germany.

## Abstract

**Background:** Anhedonia is a core symptom of major depressive disorder (MDD). While its narrow definition as a hedonic or consummatory deficit evolved to encompass anticipatory and motivational reward facets, it remains unclear where reward deficits manifest. As evidence accumulates for metabolic hormones affecting reward processing, studying their role in mitigating reward deficits could yield crucial insights. Here, we compare food reward ratings between patients with MDD and healthy control participants (HCPs) from anticipation to consummation and evaluate associations with anhedonia and metabolic parameters.

**Methods:** We conducted a cross-sectional study with 103 participants, including 52 patients with MDD and 51 HCPs. After overnight fasting, blood samples were collected to determine levels of ghrelin, glucose, insulin, and triglycerides. Participants completed a taste test, providing repeated ratings of wanting and liking, gradually moving from reward anticipation to consummation.

**Findings:** Patients with MDD showed decreased wanting (*p* = .046) but not liking for food rewards during visual anticipation. However, once food was inspected and tasted, patients increased wanting relative to HCPs (*p* = .004), providing strong evidence against a consummatory deficit (Bayes Factors > 9). In contrast to a narrow definition of anhedonia, higher scores on the Snaith-Hamilton Pleasure Scale were associated with reduced anticipatory food wanting (*p* = .010) and more pronounced increases in wanting with reward proximity (*p* = .037). Acyl ghrelin was associated with higher food reward ratings, while poor glycemic control was linked to symptoms of anhedonia.

**Interpretation:** Our study demonstrates that MDD and anhedonia are associated with reduced anticipation of rewards rather than consummatory pleasure deficits. Notably, ghrelin’s association with elevated reward ratings implicates the gut-brain axis as a potential target for treating reward deficits in MDD.

**Funding:** DFG KR 4555/7-1, KR 4555/9-1, KR 4555/10-1, and & WA 2673/15-1

**Graphical Abstract:** 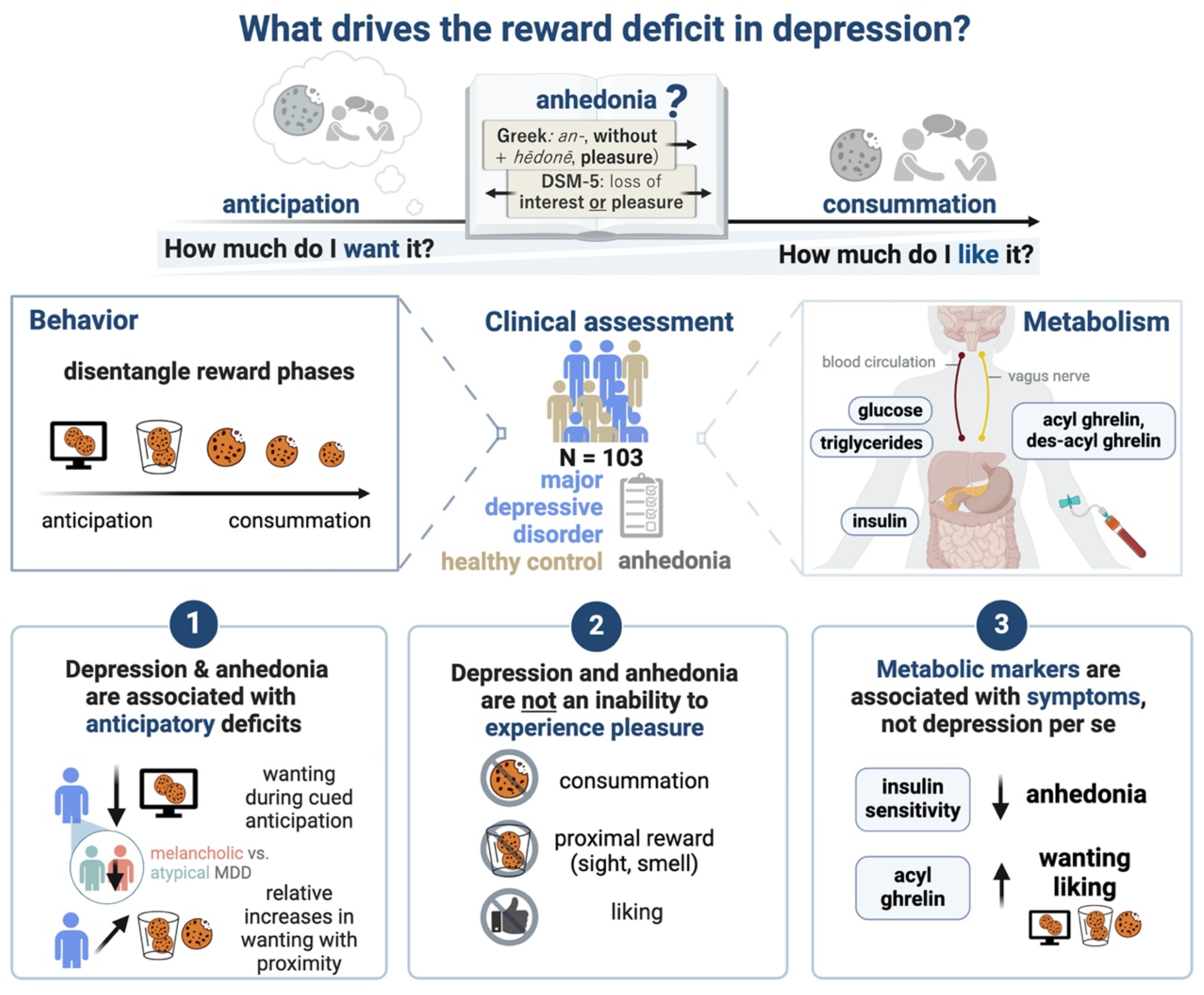

**Research in context:** *Evidence before this study:* Anhedonia, a core symptom of depression, has traditionally been conceptualised as a deficit in consummatory pleasure. However, recent definitions have expanded to include anticipatory and motivational aspects of reward processing. Despite this evolution, experimental studies that systematically investigate these facets are lacking, with most animal models of anhedonia focusing on consummatory deficits.

*Added value of this study:* This study contributes novel insights by demonstrating that patients with Major Depressive Disorder (MDD) and anhedonia exhibit reduced anticipatory wanting but not consummatory liking for food rewards. Bayesian hypothesis testing indicates strong evidence against the hypothesis of a consummatory deficit, suggesting instead a primary impairment in reward anticipation in anhedonia. Additionally, the study highlights the association between peripheral metabolic hormones and specific aspects of reward function, shedding light on the underlying mechanisms of anhedonia. Notably, lower insulin sensitivity and higher glucose levels were linked to symptoms of anhedonia, while elevated fasting acyl ghrelin levels were associated with increased food reward ratings.

*Implications of all the available evidence:* The findings suggest that deficits in anticipatory wanting, rather than consummatory pleasure, may underlie anhedonia in MDD. This distinction has important implications for treatment strategies, as targeting incentive motivation and reward anticipation could be more effective in alleviating anhedonia symptoms. The observation that wanting improves with the proximity of the reward indicates a potential therapeutic approach focusing on enhancing reward proximity. Furthermore, the association between ghrelin and reward ratings suggests a role for gut-brain signalling in motivational symptoms, particularly in cases of depression involving loss of appetite and weight.

## 1. Introduction

As a core symptom of major depressive disorder (MDD), anhedonia is linked to worse treatment outcomes and reduced quality of life, presenting an unmet challenge for therapies (1,2). Recently, the narrow definition of anhedonia as the “decreased subjective experience of pleasure” (3) has evolved towards parsing anhedonia into different facets of reward processing, including anticipation and consummation (4–6). In translational research, reward is further dissociated into wanting (i.e., the motivational drive to pursue rewards dominating during anticipation) and liking (i.e., the hedonic pleasure derived from experiencing rewards dominating during consummation) (7,8). While ample evidence associates depression with deficits in reward processing, comprehensive investigations into anhedonia which distinguish “when” (anticipation vs. consummation) and “how” (wanting vs. liking) potential deficits manifest are scarce (8,9). Moreover, patients with MDD often experience opposing changes in appetite echoed in the reward circuit’s functional architecture, suggesting the need to investigate reward deficits concerning symptoms rather than diagnosis (10). In addition to somatic symptoms in MDD, comorbid metabolic disorders (11,12) suggest a potential modulatory role of metabolic hormones on reward processing (13–16). However, to harness the potential of metabolic hormones to alleviate reward deficits, a detailed mechanistic understanding of where reward alterations manifest is needed (17).

Preclinical research has predominantly investigated anhedonia through consummatory reward responses (i.e., hedonic capacity), although the translation of taste-related tests to human research produced inconsistent results. Seminal research found that rats consumed lower amounts of sucrose and saccharose following chronic stress exposure, mimicking the appetite loss observed in (melancholic) depression (18). Since then, preclinical studies have primarily used sucrose intake or sucrose preference tests to assess anhedonia, in which a decreased preference for sucrose is interpreted as reduced liking (19–21). However, conclusive evidence for lower pleasantness ratings of sweet solutions or deficits in gustatory or olfactory function in patients with MDD is lacking (22–27). Instead, emerging evidence suggests a role of motivation (28), decreased coupling of liking and wanting (29), or reduced reward learning (30) in anhedonia. Indeed, a recent computational analysis of the sucrose preference test has identified the contribution of several of these reward facets (often uncontrolled), such as wanting to sucrose preference tests in addition to consummatory liking (21), potentially underlying heterogeneous findings. Thus, there is a great demand to dissect reward behaviour into subcomponents beyond consummatory liking, ultimately allowing targeted interventions to normalise aberrant reward-related behaviour.

Metabolic hormones, such as ghrelin and insulin, play a significant role in reward processing, transcending their role in homeostatic food control (31,32). During fasting, more ghrelin is released, increasing food intake and incentive motivation (33) via hypothalamic action and possibly vagal projections (34,35). In support of this role, ghrelin has been linked to enhanced food cue reactivity (36,37), food odour conditioning (38), alcohol self-administration and craving (39,40), but not food palatability or consummatory reward responses (41,42). However, as our group showed previously, ghrelin has been mostly investigated with respect to anticipation in humans and rarely contrasting liking and wanting (17). Preclinical work further demonstrates that ghrelin amplifies dopamine signalling in the mesocorticolimbic circuit (31,43–47). Nevertheless, investigations into plasma ghrelin levels in depression have yielded inconsistent results (48–53). In part, such inconsistencies may stem from the heterogeneity of depressive symptoms (54). For instance, differences in metabolic dysregulation have been reported between melancholic and atypical depression (55,56) and an immune-metabolic subtype of depression for ‘atypical/energy-related symptoms’ has been suggested (57,58). Atypical symptoms might include weight gain, appetite increase, carbohydrate craving or eating, hypersomnia, fatigability, mood or energy dips in the afternoon, and social withdrawal (59). In contrast to ghrelin, insulin increases postprandially, reduces food intake (60,61), and reduces dopamine signalling (62–64). Consequently, intranasal insulin application reduces food preferences, with lower insulin sensitivity attenuating this effect (65). Likewise, diminished insulin sensitivity not only weakens the translation of hunger into motivation for rewards (66) but also serves as an indicator of the efficacy of insulin, as evidenced by its association with the signal-to-noise ratio (i.e., an indicator of the signal effectiveness) of food reward signals in the nucleus accumbens (67,68). Consistent with a metabolic subtype, lower insulin sensitivity has been proposed to contribute to atypical depression (69,70). While MDD frequently occurs with type 2 diabetes (71), it has also been linked to low insulin sensitivity in non-diabetic samples from cross-sectional studies (72,73), as well as to metabolic disturbances like elevated triglycerides and increased fasting glucose (74). Taken together, metabolic hormones modulate reward processing, with ghrelin potentially enhancing and insulin sensitivity reducing reward responses.

Here, we use a taste test design, allowing us to capture food reward ratings moving from anticipation to consummation and compare reward ratings in patients with MDD and healthy control participants (HCPs). Moreover, we evaluate associations with anhedonia and metabolic parameters. To this end, we integrate behavioural, clinical, and metabolic assessments. Specifically, we measured fasting hormone levels (serum/plasma) of participants with and without MDD, followed by repeated ratings of wanting and liking before and after tasting food snacks, gradually moving from food reward anticipation to consummation. Our hypotheses were threefold: First, we expected that participants with MDD (vs. HCPs) would report lower liking or wanting ratings during anticipation and consummation. Second, we expected higher SHAPS scores, as a measure of the hedonic capacity, to be associated with reduced liking but not wanting ratings during consummation relative to anticipation. Third, considering the potential roles of ghrelin in driving incentive motivation and insulin sensitivity in reducing food value signals, we expected heightened reward ratings with higher levels of ghrelin and lower reward ratings with insulin sensitivity. We find that depression and anhedonia are characterised by anticipatory but not consummatory deficits, while metabolic hormones are linked to specific symptoms and behaviour, not depression itself. This challenges the persistent notion of anhedonia as a deficit in hedonic capacity and underscores the potential of investigating the gut-brain axis as a target to treat motivational deficits.

## 2. Methods

This cross-sectional study characterised patients with MDD and HCPs with respect to their subjective ratings of food reward during different reward phases (i.e., from anticipation to consummation). A sample size of 50 healthy control participants and 50 MDD patients was calculated beforehand to achieve a high sensitivity (at a power 1-β = .80) for moderately sized group differences (d = .57, generic r = .27).

The sample consisted of 103 participants, matched for age and body mass index (BMI; *M_Age_* = 29.3 ± 7.3 years, *M_BMI_* = 23.6 ± 3.3 kg/m^2^ [means ± SD]; Table 1) from an ongoing study on the gut-brain axis in depression (https://clinicaltrials.gov/study/NCT05318924; (75)), including 52 participants with MDD and 51 HCPs, who had never experienced a depressive episode. Sex was recorded using self-reports. All individuals interested in participating were screened for eligibility by telephone (flowchart SI1). Individuals were included if they (1) were between 20 and 50 years old, (2) had a body mass index (BMI) between 18.5 kg/m² and 30.0 kg/m². They were excluded if they (1) ever met criteria for schizophrenia, bipolar disorder, severe substance dependence or neurological condition, or for HCP, mood or anxiety disorders, (2) met criteria for eating disorders, obsessive-compulsive disorder, trauma, and stressor-related disorder, or somatic symptom disorder within the last 12-months, (3) took medication (except anti-depressive medication for MDD), or suffered from illnesses that influenced body weight, (4) for female individuals if they were pregnant or nursing at the time. For the MDD group, individuals needed to fulfil DSM-5 criteria for MDD at screening. Individuals with comorbid anxiety disorders were also included due to the high comorbidity (76) (participant comorbidities SI2). To improve generalizability, we imposed no restrictions on treatment type (e.g., psychotherapy, pharmacological, or apps) during the recruitment. However, to minimise confounding effects due to pharmacological changes, we required patients to be on stable medication for at least two months before study participation. Individuals were recruited using flyers and advertisements on social media (Facebook, Instagram) within the area surrounding Tübingen. Before inclusion, all individuals signed written informed consent. All procedures were approved by the local Ethics Committee of the University of Tübingen, Faculty of Medicine (662/2018BO1), in accordance with the Declaration of Helsinki (as revised in 2008). The compensation consisted of money and food rewards that could be acquired through the tasks (i.e., for full completion of the study, either €50 or 5 credit points + performance-based rewards). The study took place at the Department of Psychiatry and Psychotherapy in Tübingen.

**Table 1.** Participant descriptives.

| Characteristic | Overall<br>N = 103 | HCP<br>N = 51 | MDD<br>N = 52 | Test<br>Statistic | p-<br>value<sup>2</sup> |
| --- | --- | --- | --- | --- | --- |
| **Sex, n (%)** |  |  |  | 0.78 | 0.38 |
| male | 42 (41) | 23 (45) | 19 (37) |  |  |
| female | 61 (59) | 28 (55) | 33 (63) |  |  |
| **Age [years]** | 29 .3 (7.3) | 30.5 (7.2) | 28.1 (7.4) | 1.60 | 0.11 |
| **Body mass index [kg/m<sup>2</sup>]** | 23.64 (3.25) | 23.78 (3.03) | 23.50 (3.48) | 0.45 | 0.66 |
| **Acyl ghrelin [pg/mol]<sup>1</sup>** | 174 (207) | 182 (205) | 166 (210) | 0.38 | 0.71 |
| **Des-acyl ghrelin [pg/mol]<sup>1</sup>** | 188 (104) | 187 (94) | 190 (112) | -0.15 | 0.88 |
| **Glucose [mg/dl]** | 84 (8) | 83 (7) | 85 (9) | -1.20 | 0.22 |
| **Insulin [mg/dl]** | 62 (38) | 55 (28) | 69 (45) | -1.90 | 0.063 |
| **HOMA-IR** | 1.89 (1.27) | 1.65 (0.94) | 2.13 (1.49) | -1.90 | 0.055 |
| **Triglycerides [mg/dl]** | 103 (70) | 98 (64) | 107 (76) | -0.59 | 0.56 |
| **TyG** | 4.44 (0.29) | 4.42 (0.28) | 4.46 (0.29) | -0.64 | 0.52 |
| **BDI** | 15.5 (14.4) | 3.3 (4.1) | 27.6 (9.9) | -16.00 | **<0.001** |
| **SHAPS** | 12.3 (8.3) | 6.6 (6.0) | 18.0 (6.1) | -9.50 | **<0.001** |
| **Anti-depressive medication, n (%)** |  |  |  |  |  |
| None | 79 (77) | 51 (100) | 28 (54) |  |  |
| Other | 11 (11) | 0 (0) | 11 (21) |  |  |
| SSRI | 13 (13) | 0 (0) | 13 (25) |  |  |

<sup>1</sup> Values of acyl and des-acyl ghrelin refer to data of 97 participants. Data of 5 HCPs and 1 MDD were missing. <sup>2</sup> Pearson's Chi-squared test; Welch Two Sample t-test.
Data are means ± SD if not indicated otherwise. *Abbreviations:* HCP = healthy control participants, MDD = major depressive disorder, HOMA-IR = homeostasis model assessment of insulin resistance, TyG = Triglyceride-glucose Index, BDI = Beck's Depression Inventory, SHAPS = Snaith-Hamilton Pleasure Scale, SSRI = Selective serotonin reuptake inhibitors.

### 2.1. Procedure

#### 2.1.1. Experimental procedure

Participants were invited to the laboratory for two parts: a clinical interview and a behavioural intake session. Due to the different durations of the clinical interview, HCPs usually completed both parts on one day, while participants with MDD completed them on separate days. During the first part, all participants completed the Structured Clinical Interview for DSM-V (SCID-5-CV; First, 2015; ∼1.5–2 h for MDD, ∼30 min for HCP). In addition, participants with MDD completed the Structured Interview Guide for the Hamilton Rating Depression Scale with Atypical Depression Supplement (SIGH-ADS; Williams & Terman, 2003). The second part included fasting blood draws and a battery of reward-related tasks on the laptop (∼3.5 h). After a 12 h overnight fast – during which participants were instructed only to consume unsweetened beverages (e.g., water or coffee), participants answered state-related questions on a visual analogue scale (VAS) repeatedly to indicate their current subjective metabolic (i.e., feelings of hunger, fullness, and satiety) and affective state. Blood samples for the determination of acyl and des-acyl ghrelin (EDTA plasma), glucose (fluoride EDTA plasma), insulin (serum), and triglycerides (lithium heparinised plasma) were taken upon arrival by using Monovettes (Sarstedt, Nümbrecht, Germany). Afterwards, information was recorded on the participant’s last meal and drink, on anthropometric data (e.g., body weight and height), and, in the case of female participants, on the menstrual cycle phase (75). Then, participants started with a battery of reward-related tasks. As part of this battery, they completed a food cue rating task (∼15 min; (78))) and, ∼ 50 min later, a taste test (∼25 min). During the study session, individuals were provided with water ad libitum. The session concluded with participants receiving their financial compensation.

### 2.2.2. Measures

#### 2.2.1. Hormone levels

Monovettes were transferred to the Central Laboratory of the Institute of Clinical Chemistry and Pathobiochemistry of the University Hospital Tübingen for analysis of glucose, insulin, and triglycerides in plasma or serum. Glucose was determined in sodium fluoride plasma using an enzymatic test kit (Atellica CH Glucose Hexokinase_3; Atellica Solution, CI analyser), insulin in serum using an immunological assay (Atellica IM Insulin; Atellica Solution, IM Analyzer) and triglycerides in lithium heparin plasma using of an enzymatic assay (Atellica CH Triglycerides_2, Atellica Solution, CI Analyzer; Siemens Healthineers, Eschborn, Germany; within-laboratory precision for glucose ≤ 2.2%, for insulin ≤ 10%, and for triglycerides ≤ 4.0% according to manufacturer). Plasma samples for analysis of ghrelin were obtained from K3E-EDTA Monovettes immediately by centrifugation of the blood samples at 4°C with 2000 g for 10 min. Then, 500 µl of plasma was transferred into two cooled cryo tubes (Thermo Scientific™ Nunc™) each and 50 µl of cooled 1 M hydrochloric acid (HCl) in plasma to acid ratio of 10:1 was added to each tube to prevent ghrelin from deacetylating. The tubes were immediately capped, gently reversed, and cooled at - 20°C before they were stored at −80°C (after 24 to 48 h). After completing the trial, the frozen samples were transferred on dry ice to the University of Bonn. The concentration of both acylated and unacetylated ghrelin was determined by using ELISA kits (#A05306 and #A05319; both from Bertin Bioreagent, Bertin Technologies, Montigny-le-Bretonneux, France; distributed by BioCat, Germany) at the Institute of Nutritional and Food Sciences, Human Nutrition.

#### 2.2.2. Food cue reactivity and taste test

To assess different facets of reward processing, we used a food cue reactivity (FCR) task (distal sight; (78)) and a taste test paradigm (proximal sight/smell and tasting; Fig.1a). Participants rated how much they liked and wanted 7 snacks during 5 phases (1^st^ food cues, 2^nd^ proximal inspection of snack and smelling the actual snack, 3^rd^ – 5^th^ repeatedly tasting the snacks). The FCR task is a widely used task to assess food anticipation (79,80). Here, it included the 7 snacks of the taste test among a set of 60 food and 20 non-food images (81) optimised for visual characteristics (homogenous plate with grey background). Participants were presented with each item for 2 seconds twice before they rated them using a joystick on an Xbox controller and confirming by pressing the A button (82). The rating scale was presented for a maximum of 2.8 s. In separate trials, they rated how much they liked and wanted the items on psychophysically validated labelled magnitude scales (83). For liking, participants were asked to compare to all experienced sensations on a vertically labeled hedonic (visual analog) scale. Liking ratings ranged from −100 (strongest disliking imaginable) to +100 (strongest liking imaginable; (83). Wanting ratings were acquired using a horizontal scale and ranged from 0 (not wanted at all) to 100 (strongly wanted). The order of stimulus presentation and rating was pseudo-randomized.

The taste test included 7 snacks that were repeatedly rated during phase 2-5. The snacks were placed into separate glasses arranged in a circle on a wooden turntable. The glasses were prepared with enough material to have a few items each round. However, participants could decide for themselves how much they needed to make a reward rating, which was typically one item. Water for rinsing between trials was provided. In addition, the corresponding pictures of the FCR set were shown on a laptop screen. Analogous to the FCR, participants then rated the items regarding food liking and wanting. They also rated the snack’s intensity, sweetness, saltiness, and savoriness (85). As snacks were used pretzels (399 kcal/100g), NicNac’s (555 kcal/100 g; 527kcal/100 g for the vegan alternative), and bread rings (460 kcal/100 g) as salty snacks, rice crackers (380 kcal/100 g) as neutral snack, and raisins (318 kcal/100 g), chocolate chip cookies (502 kcal/100 g; 491 kcal/100 g for vegan alternative), and strawberry gummies (354 kcal/100 g) as sweet snacks.

This design allows for adjustments to reward ratings with repeated exposure. There is substantial evidence from rodent studies and human computational work that liking and wanting are distinct concepts that should be studied in concert instead of assuming a unidimensional reward representation (7,86) and might act on different timescales. While liking might serve as an initial and editable estimate about the true, long-run worth of goods, wanting might reflect the true, long-run worth of goods (86). Therefore, we measure both, liking and wanting and analyse them as distinct yet interdependent constructs.

#### 2.2.3. Questionnaires

##### Anhedonia

To measure symptoms of anhedonia, we used the German version of the Snaith-Hamilton Pleasure Scale (SHAPS (87,88)), which is widely recognised as a measure of hedonic capacity and has been validated to measure anhedonia in clinical and research settings (89). Participants indicated on 14 items how much they agreed or disagreed (Likert scale with 4 categories) with statements about experiencing pleasure over the last few days. The statements cover interests (e.g., “I would find pleasure in my hobbies and pastime”), social interactions (e.g., “I would enjoy seeing other people’s smiling faces”), sensory experiences (e.g., “I would enjoy a warm bath or refreshing shower”), and food (e.g., "I would be able to enjoy my favourite meal"). We calculated an overall sum score ranging from 0 (minimum, no anhedonia) to 42 points (maximum, anhedonia).

##### Beck’s Depression Inventory II (BDI-II)

The BDI-II is a well-validated and widely used self-report questionnaire to assess the severity of affective and somatic symptoms of depression in clinical and research settings (21 items; (90,91)). The sum of four items (loss of pleasure, loss of interest, loss of energy, and loss of sexual interest in sex) has been used to describe anhedonia (92).

##### Atypical depression

To measure the extent of atypical depression, we calculated the atypical balance score from the SIGH-ADS (59). The atypical balance score weights the atypical items (weight gain, appetite increase, increased eating, carbohydrate craving or eating, hypersomnia, fatigability, mood or energy dips, and social withdrawal; SI3) against overall symptom presence, thus representing the percentage of atypicality, considering symptom severity, ranging from 0 (minimum) to 100% (maximum).

### 2.3. Data analysis

#### 2.3.2. Preprocessing

As a measure of insulin resistance, we calculated the homeostasis model assessment of insulin resistance (HOMA-IR) using fasting glucose and insulin levels ((93), insulin [pmol/l] /6,945) * glucose [mg/dl] / 405) as well as the triglyceride–glucose (TyG) index using fasting triglyceride and glucose levels ((94); Ln(triglycerides [mg/dl] * glucose [mg/dl]) / 2). Since the distribution of hormone levels was skewed, data were log-transformed for parametric analyses, a common tool for biological data (36,95). Likewise, we log-transformed the HOMA-IR (96), whereas the TyG is already log-transformed per definition (94). We checked that log transformations resulted in approximately normal distributed values (SI4). To control for potential confounding effects of sex, age, and BMI, hormone values were residualised for these variables before being entered into the models (97,98). By using residuals, variance attributable to the linear effects of sex, age, and BMI is accounted for, ensuring that the observed hormone effects are incremental to such participant characteristics. Additionally, sex, age, and BMI were included as nuisance regressors in all models to control for their potential effect on the outcomes. While our study was not powered to test sex-specific interactions with, we report significant results for these nuisance regressors to inform future work.

### 2.4. Statistics

#### 2.4.2. Primary hypotheses

##### Linear Mixed Effects Modeling for repeated food reward ratings

For our first hypothesis of reduced wanting and/or liking ratings in MDD, we used two non-independent linear mixed-effects models using restricted maximum likelihood estimation (SI5; Model 1-2). Specifically, we modelled the dependent variables (wanting and liking) as outcome, using the following independent variables as predictors: Group (dummy coded, 0: HCP, 1: MDD), Phase (dummy coded, 0: cued anticipation), and their interaction Group x Phase to test for group differences in relative changes between reward phases. Furthermore, we included Snack type (sum coded), and all models included BMI, age, and sex (centred) as potential confounders. To account for inter-individual variance in repeated ratings, we included random intercepts and slopes for snack type and phase. For our second hypothesis of altered liking but wanting ratings in anhedonia, we replaced Group with SHAPS (centred; SI5; Model 3-4). To decide how to model the factor phase that captures ratings from anticipation to consummation we used model comparison (SI6, Fig. 1B). We refer to the terminal stage of motivated behaviour as the consummatory phase, following the definition provided by Salamone and Correa (84). This term does not refer to the act of consumption but rather to ‘consummation,’ meaning ‘to complete’ or ‘to finish’ the motivational stage, reflecting the direct interaction with the goal stimulus. Therefore, we compared three models, capturing phase as (a) a 2-level dummy factor with phase 1 + 2 (food cues, and proximal sight/smell) vs. phase 3-5 (tasting); (b) a 3-level dummy factor with phase 1 (food cues), phase 2 (proximal sight/smell, and phase 3-5 (tasting); and (c) as a 2-level dummy factor with phase 1 (food cues) and phase 2-5 (proximal sight/smell) and tasting). Model (c) outperformed the initial model (SI6, b-d). In line with the definition by Salamone and Correa (84) we refer to phase 1 as anticipation and phase 2-5 as consummation. Importantly, we report that results do not differ qualitatively depending on phase coding (SI11).

**Figure 1:**
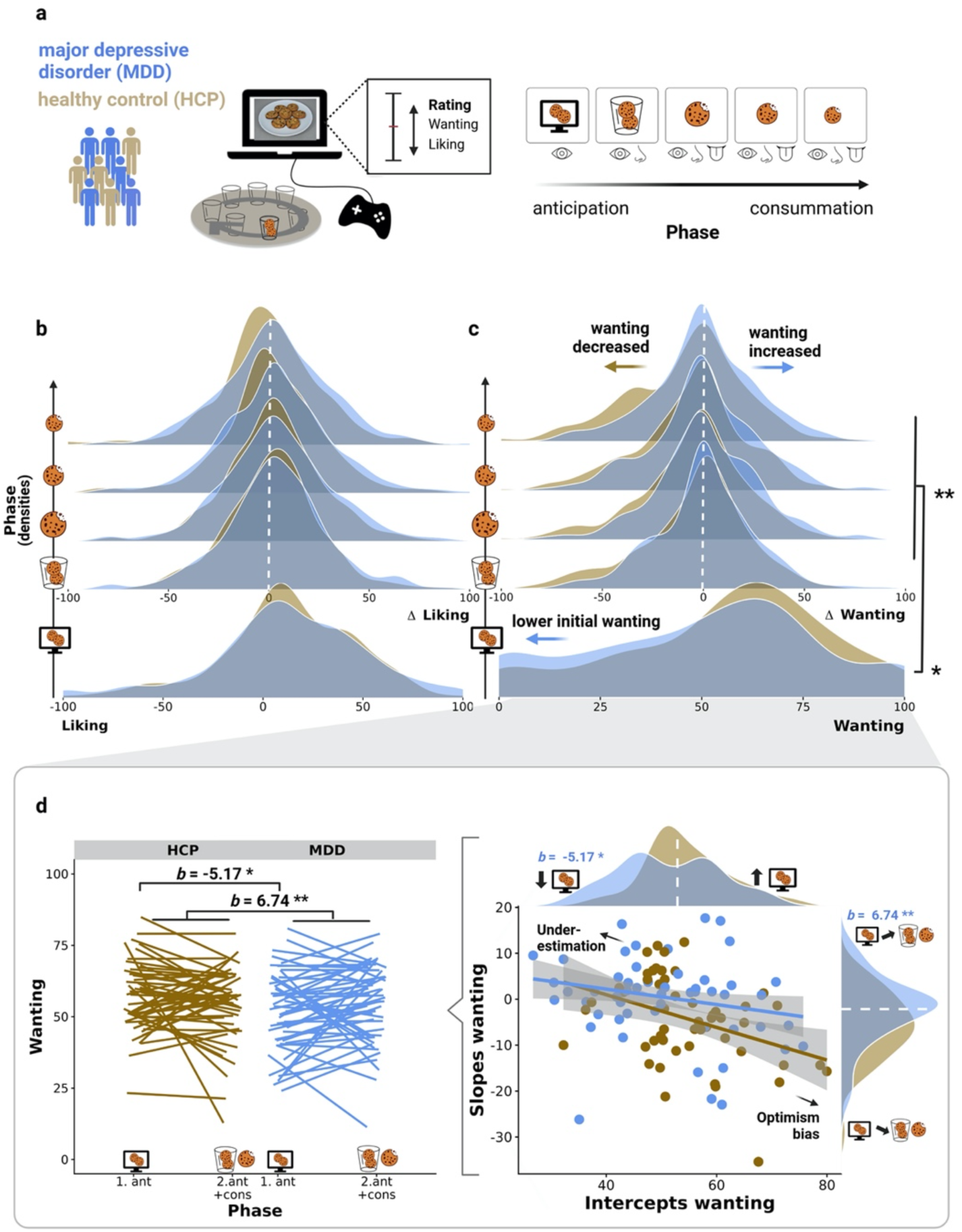
Patients with major depressive disorder (MDD) showed attenuated wanting but not liking during the anticipation of distant food rewards. A: To disentangle reward facets, participants with MDD and healthy control participants (HCP) repeatedly rated liking and wanting of snacks, moving from anticipation to consummation. B: No group differences in liking ratings (*b* = 0.21, [CI: −6.54; 6.96], *p* = .95), even after tasting food (*b* = 2.87, [CI: −2.10; 7.84], *p* = .26). C: Participants with MDD showed lower wanting ratings during anticipation (*b* = −5.17, [CI: −10.10; −0.15], *p* = .046). Once the food was proximal, wanting ratings aligned as patients with MDD reported increases in wanting compared to HCPs (*b* = 6.74, [CI: 2.19; 11.28], *p* = .004). D: Individuals with higher initial wanting ratings tended to decrease their ratings with proximal food, reminiscent of an optimism bias in HCPs. Conversely, individuals with lower initial wanting ratings tended to increase their ratings with proximal food, resembling an underestimation bias in MDD. Depicted are individual regression lines (left) and the correlation of unbiased (i.e., not including group) random intercepts and slopes derived from mixed-effects models, where differences in wanting ratings are predicted with phase and snack item (right). * *p* < .05, ** *p* < .01. Created with BioRender.com.

##### Multivariate tests for associations of metabolic hormones and reward ratings

For our third hypothesis, we used multivariate regression to evaluate potential associations of liking and wanting ratings with ghrelin and insulin sensitivity while accounting for Group and Phase and the interaction of Group and phase (SI5).

##### Bayesian analyses

Using Frequentist statistics, we can state whether data is unlikely to be observed given the null hypothesis. Based on p-values that quantify this, we can then reject or accept an alternative hypothesis (i.e., Neyman Pearson approach). However, we cannot quantify the support for the null hypothesis or a more continuous support for the alternative hypothesis (99). In contrast, Bayesian testing allows us to derive an estimate of the strength of evidence for or against specific hypotheses given the likelihood (i.e., the observed data). According to Hypothesis 2, anhedonia is often conceptualised as a consummatory deficit. Given the prevalent notion in the literature that anhedonia is characterised by a “pleasure deficit,” especially in pre-clinical models, we aimed to highlight the strength of evidence our data provides against this hypothesis. The Bayesian approach can directly evaluate the relative strength of evidence for null and alternative hypotheses by providing Bayes Factors (BF). A BF_+0_ of 3 indicates that the data is 3x more likely to happen under the alternative hypothesis than the null hypothesis. To evaluate the strength of evidence in rating changes across phases between groups (MDD vs HCPs), we used Bayesian independent samples t-tests as implemented in JASP (JASP Team, 2024, v 0.18.3) using default effect size priors (Cauchy scale 0.707). Changes in ratings and their correlation with SHAPS were analysed using Bayesian correlation tests using a stretched beta prior with a width of 0.3, as large correlations have rarely been found in psychological research (88,89). Likewise, to evaluate the strength of evidence of an anticipatory deficit, we repeated these tests for anticipatory ratings for MDD vs HCPs and their association with the SHAPS.

#### 2.4.3. Exploratory analyses

##### Depression subtype

To assess the potential influence of depression subtype and to inform future studies, we also report the results when including depression subtype. We stratified the MDD sample into participants with low atypical MDD (below median atypical balance score) versus high atypical MDD (above median atypical balance score), allowing to include the categorial *Atypical Group Fact*or (HCP vs. low atypical balance vs. high atypical balance) to test across the whole sample.

##### Hormonal values

For testing the associations of metabolic hormones with MDD and anhedonia, we used linear models (SI5).

##### Coupling of liking, wanting, and ghrelin

Since ghrelin was associated with liking and wanting, we tested whether it influences the association between liking and wanting (SI5).

#### 2.4.4. Sensitivity analyses

##### Medication

To assess the potential impact of medication, we also report how including medication type influences results by including it in the model by grouping the MDD sample: (1) no antidepressant medication, (2) selective serotonin reuptake inhibitors (SSRIs), and (3) other antidepressant medication, using treatment coding with no antidepressant medication as baseline (Table 1).

##### Depression severity

To account for the influence of depression severity, we also report how the results of interest change when including the BDI sum score (centred).

##### Anhedonia measurement

To exclude that our results solely rely on our choice of instrument to measure anhedonia (i.e., SHAPS sum score), we also report results for single SHAPS items and results exchanging SHAPS for the BDI anhedonia subscore (centred; SI7).

##### Liking and wanting dependency

Since ratings of wanting and liking are associated (*r* = .75, [CI: .65; .82]; *p* < .001), we added liking as a predictor to the interaction of phase and MDD (or anhedonia) in the models with wanting as outcome, to isolate variance that is not captured by liking ratings (SI8)

##### Other taste ratings

Since anhedonia has been associated with differences in sweet taste perception, which could explain differences in wanting or liking we also tested whether participants with MDD and anhedonia differed in any of the taste ratings (SI15).

##### Influence of potential violation of heteroscedasticity

Since there was some indication of a violation of the heteroscedasticity assumption, we corroborated the linear mixed-effects models with the wild bootstrapping method to check the robustness of the parameter estimates. The wild bootstrap makes no distribution assumptions and allows for heteroskedasticity (100). We report a comparison of parametric and non-parametric estimates and standard errors (SI9). Importantly, our conclusions do not change qualitatively.

#### 2.4.5. Software and thresholds

Primary analyses were conducted with R (v4.3.2; R Core Team 2021). For statistical modelling, we used the *lmer* and *summary* function of the ‘*lmerTest’* package v3.1.3 (101) which estimates degrees of freedom using the Satterthwaite approximation. For multivariate regression analysis, we used the *lm* and *anova* function. For wild bootstrapping, we used the *lmeresampler* package. For Bayesian tests, we used JASP (JASP Team, 2024, v 0.18.3). For our primary hypotheses, we considered α < 0.05 as significant. Exploratory analyses (a. depression subtype, b. group differences in hormones, c. SHAPS associations with hormones) were corrected by controlling the false discovery rate (102), (SI5 for hypotheses families).

## 3. Results

### 3.1. Descriptive data

Among the 103 participants, 42 were male, and 61 were female, with a mean (SD) age of 29.3 (7.3) years and a mean (SD) body mass index (BMI) of 23.63 (3.25). HCPs and patients with MDD did not differ in age, sex, or BMI (Table 1). Patients with MDD (vs. HCPs) had higher scores on the BDI-II and anhedonia as measured by higher SHAPS scores. The groups did not differ on any of the measured (uncorrected) blood parameters. Further details on the patient population are presented in Table 1; (corrected) results taking into account age, sex, and BMI arre reported in Results 3.5.

### 3.2. Lower wanting but not liking during initial anticipation in MDD

To disentangle reward facets in depression, we developed a taste test in which participants with and without MDD repeatedly rated food liking and wanting, moving from anticipation to consummation (Fig. 1a). Using linear mixed-effect models, we modelled liking and wanting ratings to evaluate group differences throughout the taste test (full model output, SI10). Since individuals adjusted their ratings once the food was present in front of them (i.e., after an initial anticipatory rating, Fig. 1c), a model separating first anticipation (i.e., visual food cues) from consummation (i.e., proximal inspection and sequential tasting; fit the data best (model comparisons, SI6). Therefore, we used the 2-level phase factor to separate anticipation from consummation (including sight, smell, touch, and taste) for all further analysis. From a theoretical viewpoint, regarding sight and smell as part of the consummatory phase is reasonable because consummation describes the terminal stage of motivated behaviour and does not only refer to “consumption”. Instead, it means that “motivational stimuli are available at some physical or psychological distance from the organism” (84). Importantly, using the three-phase model or the reported two-phase model does not qualitatively change our conclusions (SI11).

Cookies were the preferred snacks, around 26 points higher on the liking scale and 23 points higher on the wanting scale than on average, and raisins were the least liked snacks, around 17 points lower on the liking and 14 on the wanting scale than on average. A difference of 6.2 points on the liking scale corresponds to the difference between “neutral” and “like slightly”. During the first anticipation, participants with MDD reported similar food liking (*b* = 0.21, [CI: −6.54; 6.96], *p* = .95, Fig. 1b), but lower food wanting (*b* = −5.17, [CI: −10.10; −0.15], *p* = .046; Fig. 1c) compared to HCP. During the consummatory phase, participants with MDD reported similar liking (*b* = 2.87, [CI: - 2.10; 7.84], *p* = .26) and no longer indicated lower food wanting (*b* = 1.56, CI: −3.27; 6.40], *p* = .53). Consequently, participants with MDD increased wanting compared to HCPs who decreased wanting (*b_Phase_* = −5.74, [CI: −9.04; −2.44], *p* = .0009, *b_MDDxPhase_* = 6.73, [CI: 2.19; 11.28], *p* = .004; Fig. 1d). Notably, we did not observe any sex differences or dependencies on medication type. These results indicate differences in the incentive salience of distant, not proximal rewards, and there were no differences in pleasure when tasting food in MDD.

MDD is a heterogeneous condition, and patients may experience increases or decreases in appetite and body weight during a depressive episode (SI12), with melancholic depression being characterised by decreased appetite. Accordingly, we explored whether results change using the depression subtype. Patients with melancholic MDD primarily reported blunted food wanting during anticipation (vs. HCP: *b* = −8.97, [CI: −14.95; −2.99], *p_FDR_* = .012, vs. atypical MDD: *b* = 7.77, [CI: 0.80; 14.73], *p_FDR_* = .07, Fig. S3). In contrast, patients with atypical MDD did not initially show reduced food wanting (vs HCP: *b* = −1.20, [CI: −7.30; 4.91], *p_FDR_* = .70, Fig. S3). Despite differences in initial ratings, all patients with MDD showed comparable increases in wanting during the consummatory phase (atypical: *b_GroupxPhase_* = 7.80, [CI: 2.15; 13.44], *p_FDR_* = .012 vs. melancholic MDD: *b_GroupxPhase_* = 5.73, [CI: 0.23; 11.24], *p_FDR_* = .044, Fig. S3). Consequently, patients with atypical MDD even reported higher wanting during consummation compared to HCPs, however, this did not survive control for false discovery rate (*b* = 6.60, [CI: 0.83; 12.37], *p_FDR_* = .081). In contrast, we did not find differences in liking between melancholic (*b* = −2.05, [CI: −10.22; 6.11], *p_FDR_* = .93) and atypical MDD (*b* = 2.54, [CI: −5.80; 10.88], *p_FDR_* = .93) compared to HCPs, or between depression subtypes (*b* = −4.60, [CI: −14.10; 4.91], *p_FDR_* = .62).

### 3.3. Anhedonia is not associated with reduced consummatory liking

After demonstrating reduced wanting ratings during anticipation in depression, we investigated specific associations of ratings during anticipation and consummation with anhedonia (as measured by higher SHAPS scores; full model output SI13). In contrast to the alleged reflection of impaired hedonic capacity (i.e., consummatory liking), higher SHAPS scores were associated with reduced wanting (*b* = −0.40, [CI: - 0.70; −0.10], *p* = .010, Fig. 2c), but not liking (*b* = −0.36, [CI: −0.76; 0.04], *p* = .081, Fig. 2a) during the first anticipation rating. Furthermore, participants with higher SHAPS scores showed increases in wanting ratings for proximal rewards (*b_SHAPSxPhase_* = 0.30, [CI: 0.02; 0.58], *p* = .037, Fig. 2d). In contrast, liking ratings did not change (*b_SHAPSxPhase_* = 0.25, [CI: −0.03; 0.52], *p* = .080, Fig. 2b). Single items did not drive associations of anhedonia with reduced anticipatory wanting since we observed negative coefficients for all SHAPS items (Fig. 2e). Still, taste-related items (i.e., enjoying favourite food, enjoying a favourite drink) were among the three most robust predictors for reduced anticipatory wanting. For the interaction with phase, we observed positive associations (i.e., increased wanting with proximal rewards) for all SHAPS items. However, different items showed the strongest association compared to anticipation (Fig. 2e). Older individuals (*b* = -.33, [CI: −0.64; −0.01], *p* = .047) showed overall lower wanting ratings, but this did not influence the associations with the SHAPS. Depression severity (using the BDI) did not explain lower initial wanting (*b* = −0.15, [CI: −0.33; 0.02], *p* = .09); however, severity was associated with the observed increases in wanting with proximal food (*b_BDIxPhase_* = .20, [CI: 0.03; 0.36], *p* = .018). As some items from the BDI tap into anhedonia (103), we also investigated the BDI anhedonia subscore, partially replicating the pattern for SHAPS (SI7). Since lower wanting ratings were associated with SHAPS and melancholic MDD, we inspected the correlation between SHAPS and depression subtype (*r* = -.082, *p* = .56, SI14), but these dimensions are largely orthogonal and may contribute independently to altered wanting ratings. Likewise, medication type did not alter the results. Notably, neither depression nor anhedonia was characterised by differences in perceived taste (SI15), further corroborating that depression and anhedonia are not associated with altered taste perception per se. As reported previously by our group, patients with MDD did not differ from HCPs in subjective ratings of metabolic state (i.e., hunger, fullness (75)).

**Figure 2:**
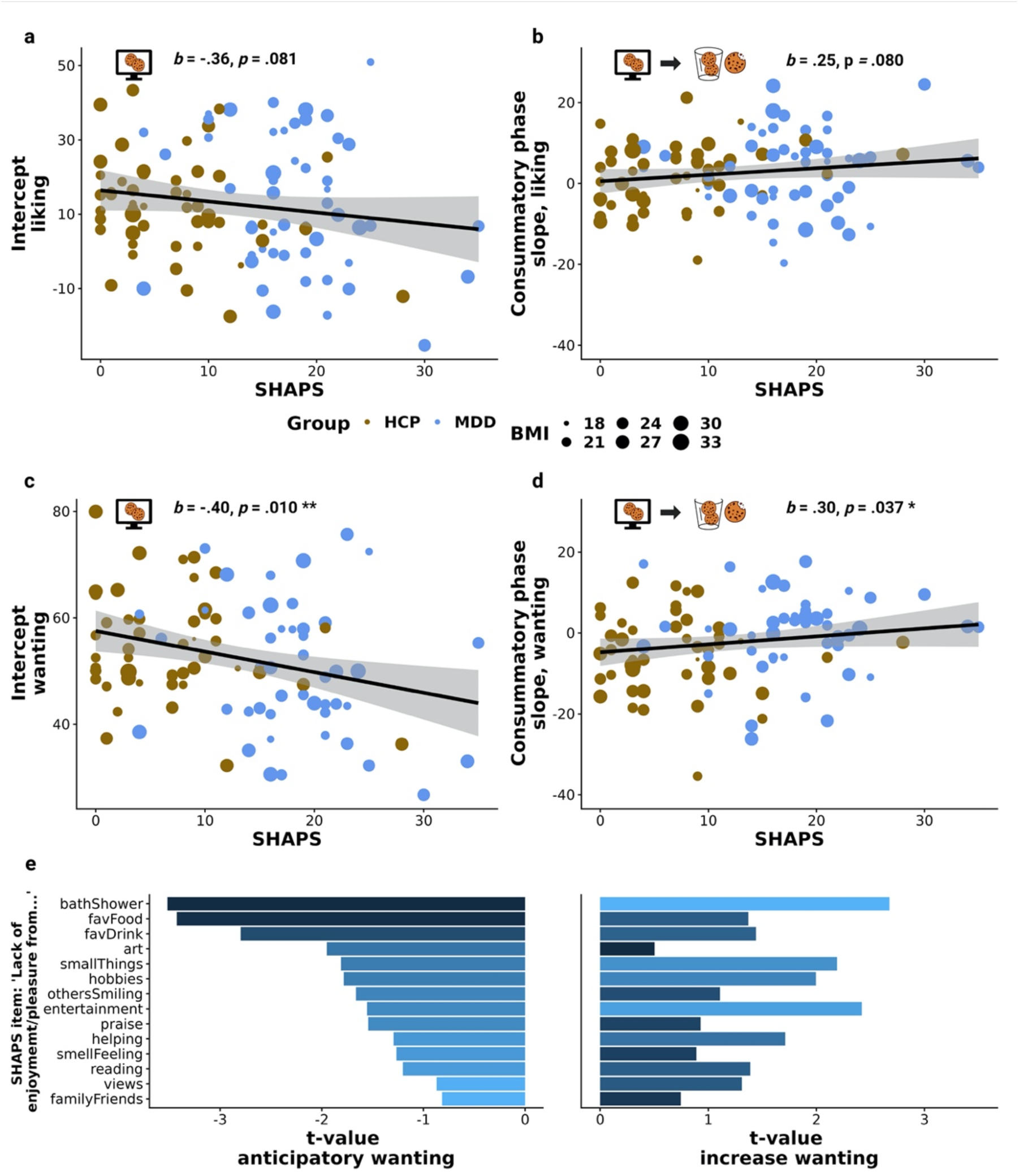
Anhedonia was associated with blunted wanting of food rewards during anticipation but increased wanting with reward exposure. A: Higher SHAPS scores were weakly associated with lower liking during cued anticipation (*b =* -.36, [CI: −0.76; 0.04], *p* = .081). B: Higher SHAPS scores were not associated with increased food liking during the consummatory phase (*b =* .25, [CI: −0.03; 0.52], *p* = .080). C: Higher SHAPS scores were associated with lower wanting during cued anticipation (*b =* -.40, [CI: −0.70; −0.10], *p* = .010). D: Once food is proximal, higher SHAPS scores were associated with increases in wanting ratings after cued anticipation (*b* = .30, [CI: 0.02; 0.58], *p* = .037). For A-D, we depicted individual intercepts derived from an unbiased mixed-effects model (i.e., not including SHAPS), where differences in ratings were predicted by phase and snack items. E: All SHAPS items were negatively associated with anticipatory wanting, with imagining taking a bath, eating one’s favourite food, or drinking one’s favourite drink being the strongest predictors (left panel). Similarly, all SHAPS items were associated with increased wanting during the consummatory phase (right panel). * *p* < .05, ** *p* < .01. Created with BioRender.com

### 3.4. Moderate to strong Bayesian evidence against a consummatory deficit in depression and anhedonia

To evaluate the strength of evidence against consummatory reward deficits in patients with MDD and anhedonia provided by our study, we calculated Bayes factors (BF) for the relative changes in ratings with consummation for patients vs. controls. The observed increases in wanting provide strong evidence against an alleged consummatory deficit in MDD (BF_0+_ = 16.27; i.e., 16x more likely to occur if we do not assume a deficit) and in association with anhedonia (BF_0+_ = 12.27; i.e., 12x more likely if we do not assume a deficit, i.e., no decrease with consummation; Fig. 3). Likewise, the absence of differences in liking changes provides moderate evidence against an alleged consummatory deficit in MDD (BF_0+_ = 9.09; i.e., 9x more likely to occur if we do not assume a deficit) and in association with anhedonia (BF_0+_ = 10.69; i.e., 11x more likely if we do not assume a deficit; Fig. 3). As wanting ratings changed in a direction opposite of expectation (i.e., increased rather than decreased with consummation), we additionally tested for an undirected effect. We found moderate (MDD) and anecdotal (SHAPS) evidence that ratings increase with consummation relative to anticipation in MDD and with higher SHAPS (SI16). Prior selection did not qualitatively change these results (Robustness checks; SI17-18). Within the MDD group, we found anecdotal evidence for differences between melancholic and atypical depression in lower wanting in melancholic MDD (BF_01_= 2.33; i.e., 2x more likely to occur if we assume differences between subtypes) and stronger increases in wanting in atypical MDD (BF_01_= 2.98; i.e., 3x more likely to occur if we assume differences between subtypes). This suggests that our findings reflect both differences between patients with MDD and HCPs as well as differences within patients with MDD in accordance with atypical symptoms (i.e., increases in appetite).

**Figure 3:**
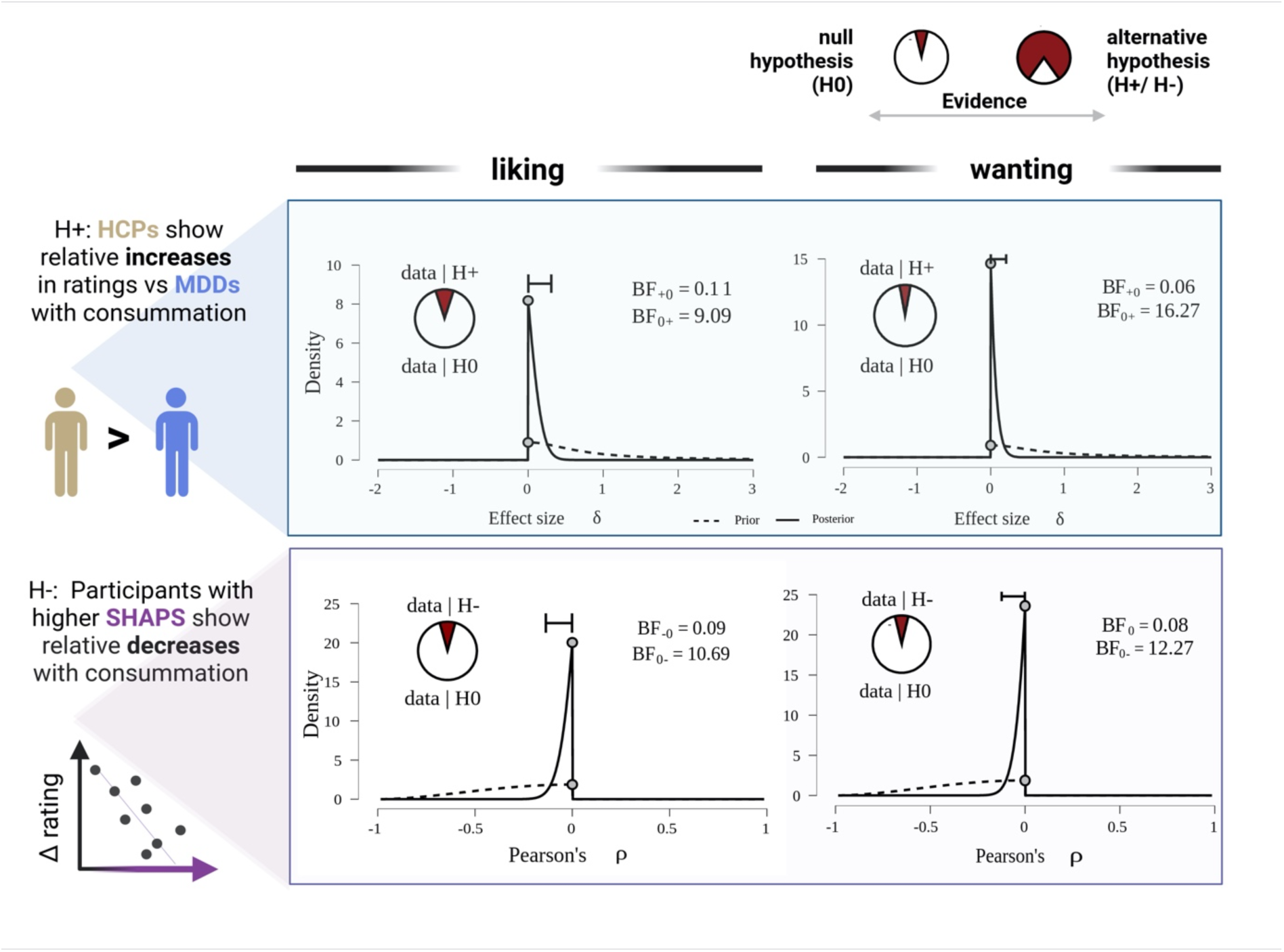
Bayesian hypothesis testing strengthens evidence against the common idea that depression or anhedonia is a consummatory deficit. Moderate to strong evidence against the common hypothesis that participants with MDD (vs HCPs) show relatively reduced liking (moderate) or wanting (strong) during consummation compared to anticipation (top panel; one-sided Bayesian independent samples *t*-test). Strong evidence against the hypothesis that higher SHAPS (i.e., lower “hedonic tone”) is associated with stronger liking or wanting decreases during consummation (bottom panel; Bayesian Negative Correlation). *BF* = Bayes factor (with levels of evidence: 1-3 anecdotal, 3-10 moderate, 10-30 strong). A probability wheel on an area of size 1 represents the BF10, respectively. Created with BioRender.com

To evaluate the strength of evidence of the observed anticipatory deficit in patients with MDD and anhedonia provided by our study, we calculated BF for absolute ratings during anticipation. The observed lower wanting during anticipation provides anecdotal evidence for an anticipatory wanting deficit in MDD (BF +0 = 2.19), and the association of lower anticipatory wanting with greater SHAPS provides strong evidence for an anticipatory wanting deficit in anhedonia (*r* = -.271; BF −0 = 17.75). The lack of lower liking in MDD compared to HCPs during anticipation provides moderate evidence in favour of the null hypothesis (BF+0 = 4.90), indicating no difference in anticipatory liking. Likewise, the non-significant association between lower liking and SHAPS provides anecdotal evidence for lower anticipatory liking in anhedonia (*r* = -.168; BF - 0 = 1.76).

### 3.5. Metabolic hormones are associated with symptoms, not MDD per se

Next, we evaluated the influence of metabolic hormones and potential disturbances on symptoms of MDD. As reported in Table 1, (untransformed) metabolic hormone values did not differ between the HCPs and patients with MDD. This held when using log-transformed values adjusted for age, sex, and BMI and when including age, sex, and BMI as covariates (Fig. 4). Fasting acyl ghrelin levels (adjusted for age, sex, and BMI) were not altered in MDD (*b* = -.27, [CI: −0.61, 0.08], *p_FDR_* = .22, exp(b) = 0.76, Fig. 4c; including depression severity: *b* = -.55, [CI: −1.18, 0.09], *p* = .094, exp(b) = 0.58). However, patients with melancholic MDD showed lower ghrelin levels compared to HCPs (*b* = -.44, [CI: −0.85, −0.02], *p* = .039, SI19, exp(b) = 0.64); however, this did not survive when controlling the false discovery rate (*p_FDR_* = .15). Likewise, ghrelin was not associated with SHAPS scores (*b* = −1.63, [CI: −3.62, 0.37], *p_FDR_* = .14, SI20). Fasting levels of des-acyl ghrelin were similar in patients with MDD and HCPs (*b* = .007, [CI: −0.20, 0.22], *p_FDR_* = .94).

**Figure 4:**
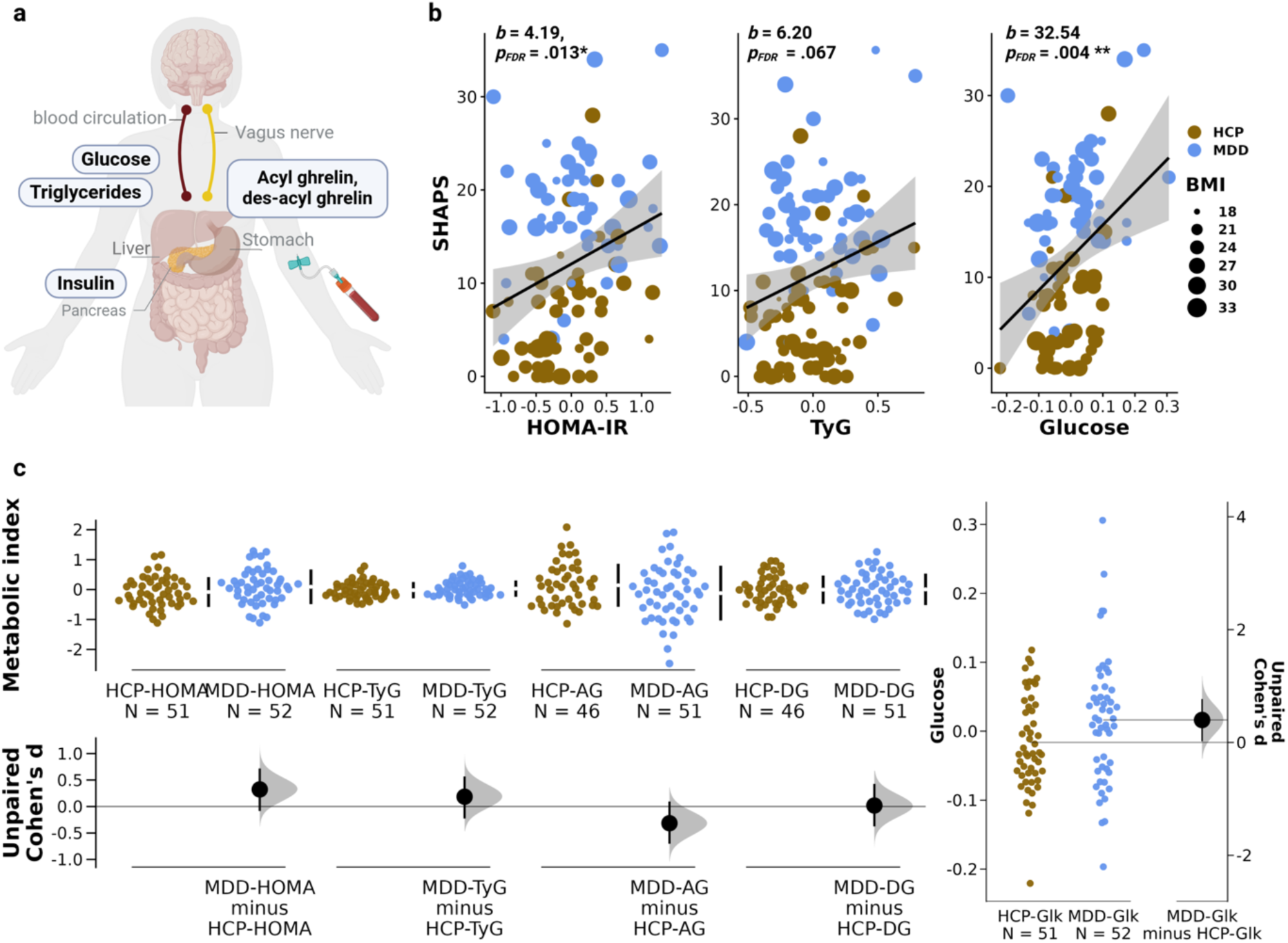
Metabolic disturbances were associated with specific symptom profiles of depression. A: Metabolic parameters were determined after a 12-hour overnight fast, including acyl and des-acyl ghrelin, insulin, glucose, and triglycerides. The latter three were used to determine two indices of insulin resistance, HOMA-IR and triglyceride index (TyG). B: Higher insulin resistance (adjusted for BMI, sex, and age) as indexed by HOMA-IR (*b* = 4.19, [CI: 1.30, 7.09], *p_FDR_* = .013) but not TyG (*b* = 6.20, [CI: 0.30, 12.11], *p_FDR_* = .067) was associated with SHAPS. Glucose levels were strongly associated with SHAPS (*b* = 33.48, [CI: 14.92, 52.06], *p_FDR_* = .004). C: Cumming estimation plots show no significant group differences in metabolic indices (HOMA-IR, TyG, Acyl- and Desacyl ghrelin), except glucose levels which were higher in MDD (*b* = .03, [CI: 0.001, 0.07], *p* = .046, but this did not hold when controlling the false discovery rate*; p_FDR_* = .22). Effect size and bootstrapped 95% CIs were plotted in addition to raw data. *Note*: All hormonal values were log-transformed and residualised for sex, age, and BMI. Created with BioRender.com

Regarding glycemic control, we observed no group differences in the TyG (*b* = .05, [CI: −0.06, 0.16], *p_FDR_* = .41, exp(b) = 1.05) and HOMA-IR (*b* = .19, [CI: −0.03, 0.40], *p_FDR_* = .22; exp(b) = 1.20) in patients with MDD compared to HCPs. This held when adding depression severity or depression subtype to the models. Still, patients with MDD showed higher fasting glucose levels (*b* = .03, [CI: 0.001, 0.07], *p* = .046; exp(b) = 1.03). However, this did not survive when controlling the false discovery rate (*p_FDR_* = .22). In contrast to MDD, participants with higher SHAPS scores showed higher fasting glucose (*b* = 33.48, [CI: 14.92, 52.06], *p_FDR_* = .004; i.e., for a 10% increase in glucose, the expected outcome for SHAPS increases by 3.19), lower insulin sensitivity (*b* = 4.19, [CI: 1.30, 7.09], *p_FDR_* = .013, Fig. 4b; i.e., for a 10% increase in HOMA-IR, the expected outcome for SHAPS increases by 0.40) but not higher TyG (*b* = 6.20, [CI: 0.30, 12.11], *p_FDR_* = .067; i.e., for a 10% increase in TyG, the expected outcome for SHAPS increases by 0.59). Sensitivity analyses showed that the association of anhedonia with HOMA-IR (*b* = 2.53, [CI: 0.35, 4.71], *p* = .023; i.e., for a 10% increase in HOMA-IR, the expected outcome for SHAPS increases by 0.24) and glucose (*b* = 20.80, [CI: 6.66, 34.94], *p* = .004; i.e., for a 10% increase in glucose, the expected outcome for SHAPS increases by 1.98) exceeded the effects of MDD since including Group in the model did not fully attenuate the associations. The association with the triglyceride index remained insignificant (*b* = 4.21, [CI: −0.15, 8.56], *p* = .058, Fig. 4b), indicative that glucose levels and insulin dominate the association between anhedonia and glycemic control. Again, depression severity or depression subtype did not qualitatively alter these associations. Notably, the association between HOMA-IR and SHAPS was attenuated in melancholic MDD (*b* = −7.11, [CI: −12.28; −1.93], *p_FDR_* = .040, SI21; i.e., for a 10% increase in HOMA-IR in melancholic MDD, the expected outcome for SHAPS is 0.67 lower). We did not observe any sex differences. These results support a link between poor glycemic control and anhedonia in depression.

### 3.6. Acyl ghrelin is associated with overall higher ratings of food reward

Next, we assessed whether metabolic hormones and potential disturbances translate to differential ratings collected during the taste test. To this end, we tested for a multivariate effect of acyl ghrelin and insulin sensitivity on liking and wanting ratings including reward phase, group, and their interaction. Acyl ghrelin was associated with higher ratings overall (Pillai’s Trace *V* = .049, *F*(2, 185) = 4.74, *p* = .010). Separate models showed associations for wanting (*t* = 2.88 *p* = .005) and liking (*t* = 2.83, *p* = .005), supporting a role of ghrelin in incentive motivation (see SI22 for separate linear mixed-effects models considering the hierarchical structure of the data corroborating these conclusions). Adding depression severity to the model did not alter these associations (Pillai’s Trace *V* = .05, *F*(2, 185) = 5.34, *p* = .006). Notably, follow-up analyses showed that higher levels of acyl ghrelin reduced the correspondence between wanting and liking ratings (*b_likingxGhrelin_* = -.23, [CI: −0.39; −0.07], *p* = .005 (uncorrected); i.e., for a 10-unit increase in liking, the expected outcome for wanting is 0.22 lower for a 10% increase in ghrelin than no change in ghrelin), pointing to a potential shift in the integration of incentive salience and hedonics, such that with higher acyl ghrelin levels less liked food rewards are wanted more (Fig. 5b). This interaction did not change when adding group and depression severity to the model. Inspecting the covariates showed that females (*b* = −4.02, *p* = .028) and older individuals (*b* = -.33, *p* = .009) showed overall reduced wanting. Follow-up analyses showed this is attenuated in older participants with higher ghrelin (age; *b* = 0.59, [CI: 0.21; 0.90], *p*= .002 (uncorrected); SI23). In contrast, fasting levels of des-acyl ghrelin showed weaker and non-significant associations with wanting and liking, and we did not observe associations between HOMA-IR with wanting and liking (SI24).

**Figure 5:**
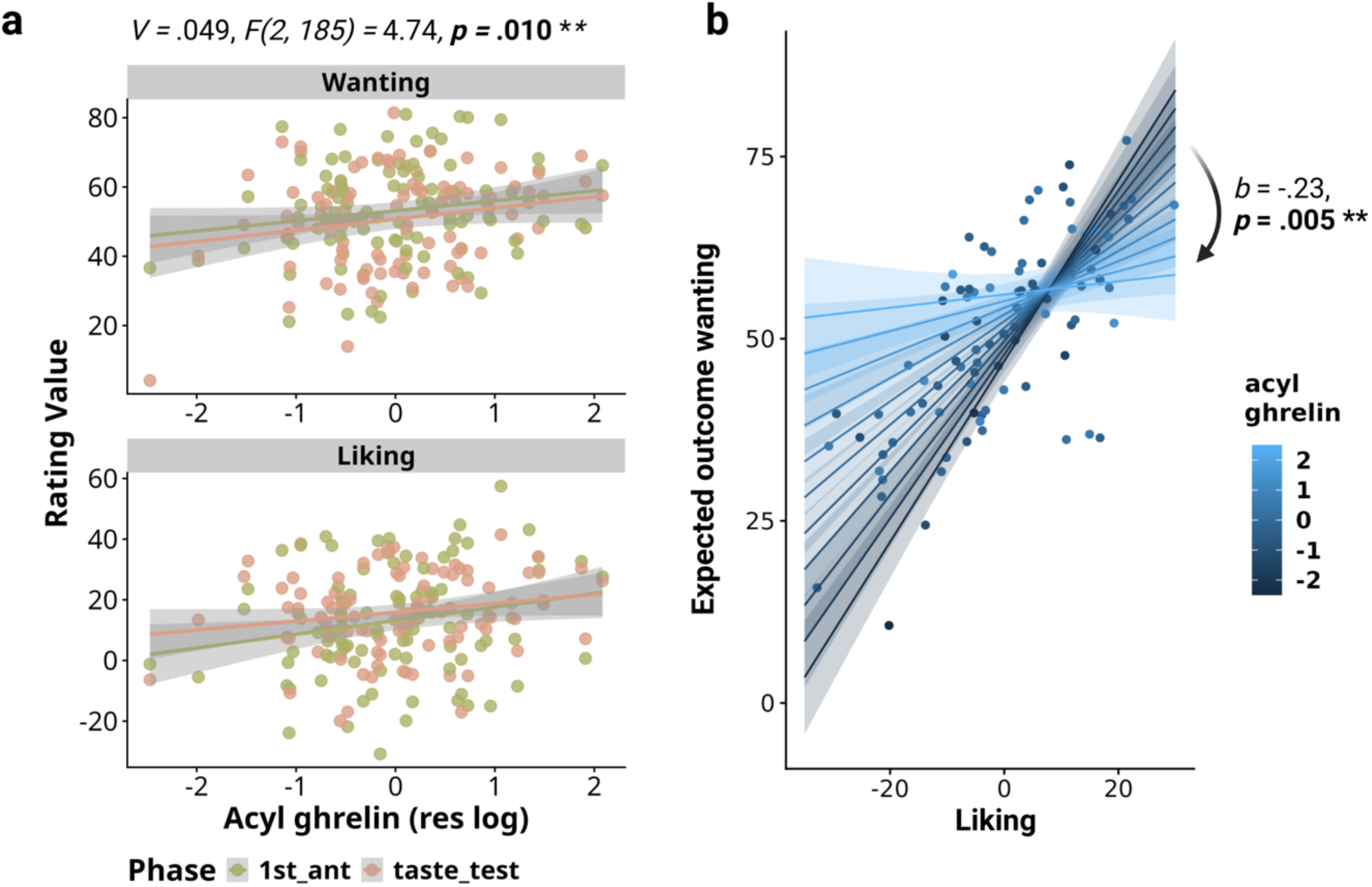
Acyl ghrelin is associated with the coupling between liking and wanting of food rewards. A: Multivariate regression showed that ghrelin was associated with greater food reward ratings and that this association was not specific to liking or wanting (Pillai’s Trace *V* = .049, *F*(2, 185) = 4.74, *p* = .010). B: Estimated marginal means of a fitted linear model to predict average wanting (across phases and snacks), using acyl ghrelin as a fixed effect and its interaction with liking ratings. Ratings of liking and wanting were strongly positively associated, but acyl ghrelin was associated with this coupling. With higher fasting ghrelin levels, food wanting became less dependent on liking (*b* = .23, *p* = .005; uncorrected). Created with BioRender.com

## 4. Discussion

An improved distinction between failing to seek pleasurable activities and not enjoying them holds actionable implications for treating anhedonia as a cardinal symptom of MDD (4,6). Here, we combined comprehensive clinical, behavioural, and metabolic assessments to localise reward dysfunction in MDD and gauge the potential for interventions targeting the gut-brain axis (13,17,31,104). First, we show that patients with MDD and anhedonia primarily experience reduced anticipatory wanting for food rewards. In contrast to the conventional notion that anhedonia is an inability to experience pleasure, we found no differences in anticipatory or consummatory liking and even relative increases in wanting during consummation. Second, our unique design shows that reward deficits are marked for distal (i.e., cued anticipation) but not proximal (i.e., sight, smell and taste) rewards, contributing to an improved mechanistic understanding of anhedonia as a motivational deficit. Third, we show that peripheral levels of metabolic hormones are associated with specific aspects of reward function rather than MDD *per se*. Lower insulin sensitivity and higher glucose levels were associated with anhedonia, whereas higher fasting acyl ghrelin levels were associated with higher wanting and liking ratings. Since ghrelin levels were lower in melancholic MDD, it is plausible that altered gut-brain signalling may contribute to motivational symptoms in melancholic MDD experiencing loss of appetite and weight. Our results corroborate the role of reward anticipation in anhedonia and highlight reward proximity and metabolic health as factors for future translational work. Crucially, increases in wanting in patients with MDD and anhedonia during reward consummation provide strong evidence against the hypothesised deficit in hedonic capacity and call for a revision of the term “anhedonia”.

Our findings extend previous work on impaired reward processing in depression by disentangling two crucial phases often investigated separately before (9): anticipation and consummation. Blunted wanting ratings during anticipation might reflect lower incentive motivation in MDD, reducing the tendency to approach a reward (‘wanting’; (105)) despite comparable ratings of pleasantness and taste quality. Subjective ratings of wanting have been linked to the recruitment of core regions within the reward circuit (106,107) and dopamine neurotransmission for food cues during anticipation (108,109), indicative of incentive motivation to pursue rewards. In depression, blunted recruitment of the reward circuitry during anticipation of incentive cues has been reported ((110–113) but (114)), supporting that the desire for rewards might be altered in MDD. This mechanism may contribute to symptoms of melancholic MDD, which has been associated with the failure to develop a biased response for more frequently rewarded stimuli (115) and deficits in reward anticipation during a slot machine task (15). Accordingly, within the MDD group, we found reduced wanting in the melancholic subtype. Crucially, anhedonia was more strongly associated with blunted wanting (vs. liking) ratings during anticipation, followed by larger increases in wanting during consummation, thereby contradicting the conventional notion that anhedonia reflects an inability to experience pleasure (4,116). As anhedonia questionnaires inherently assess the recollected experience of distant rewards, they reflect subjective representations of motivational value or negativity bias (117). Consequently, they align more closely with processes involved in cued anticipation rather than direct hedonic experiences. In support of this notion, our results argue against using questionnaires, such as the SHAPS, as measures of hedonic capacity (118). Instead, our presented results suggest that behavioural assessments provide more nuanced insights into reward deficits that may better guide future translational work than questionnaires alone.

One strength of our study design is that it resolves intra-individual changes across phases, pinpointing lower food anticipatory wanting in patients with MDD and anhedonia. In principle, two processes may explain the group differences in anticipation but not consummation: (1) overestimation of reward value during anticipation in HCP (119), or (2) underestimation of reward value during anticipation in MDD (120). Our findings support both processes: while wanting decreased in HCPs during consummation, it increased in patients with MDD. Crucially, larger corrections of an initial negative bias were associated with anhedonia, substantiating that anhedonia is primarily related to altered motivational reward anticipation (28). Our findings also argue against a mechanistic deficit in reward learning that drives anhedonia (30) as the differences in wanting ratings faded already in the mere presence of the rewards. Since beliefs about the distance to a reward or desirable state may dictate wanting and instrumental motivation (121), participants may differ in their reward-related expectations, irrespective of momentary enjoyment during consummation. For instance, internal beliefs that increase the perceived distance of a reward might reduce hedonic experiences (122). Accordingly, negative biases in patients with MDD concerning rewards have been reported before with diverse paradigms (120,123,124). Taken together, blunted reward anticipation might be explained by perceived reward distance more than by a failure to experience or learn from rewards. Increased reward proximity is targeted by behavioural activation therapy, which augments the exposure to rewards. However, behavioural activation therapy has led to heterogeneous results, possibly because it does not address the negative bias during anticipation effectively and instead capitalises on reward responsiveness (125,126). Therefore, additional refinements are necessary to treat anhedonia more effectively.

By combining precision-oriented clinical and behavioural assessments with metabolic profiling, our study illustrates the potential of metabolic hormones to modulate reward responses (15–17). In line with the comorbidity between MDD and type 2 diabetes (71), lower insulin sensitivity and higher fasting glucose levels were strongly associated with anhedonia. Likewise, hyperglycemia, diet-induced changes in insulin signaling, and knockout of insulin receptors facilitate depression and anhedonia-like behaviour in rodents (63,127–129). However, we did not observe altered metabolic hormone concentrations in MDD compared to HCPs when adjusted for age, sex, and BMI; only higher glucose levels were found in MDD, supporting that primarily the anhedonic subtype of MDD is associated with metabolic dysregulation (130,131). Although neither insulin sensitivity nor glucose levels affected food ratings, it is plausible that the effect is smaller if food is only tasted and not consumed ad libitum. In contrast to glucose, fasting levels of acyl ghrelin were not different in depression; however, taking depression subtypes into account revealed lower levels of acyl ghrelin in melancholic MDD; albeit this did not hold up correction for multiple tests. At the same time, higher ghrelin was associated with higher wanting and liking ratings across reward phases. This is in accordance with (preclinical) studies showing that ghrelin increases food cue reactivity (36,37), food intake (132), greater motivation to work for food (47), and is involved in augmenting various drug rewards (39,133,134), mainly via increased dopamine transmission in the mesocorticolimbic pathway (134,135). The association of ghrelin and subjective ratings of wanting and liking across phases of the taste test supports the role of ghrelin in increasing the ‘appetizer effect’ (136), as fasting levels of ghrelin have been associated with increases in subjective appetite during the initial stages of meals (36). Furthermore, ghrelin might modulate dopamine transmission and alter reward expectancy signals during cued rewards (137). With higher ghrelin levels, wanting ratings were less coupled with liking, indicating that during an energy deficit, the motivation for food rewards increases more independently of the hedonic impact (138). Given the conflicting evidence on ghrelin’s action in depression (70), we do not find clear evidence for lower ghrelin levels in depression. Yet, a stronger contribution of ghrelin to heightened reward function might be of high clinical relevance for melancholic MDD because this group showed the anticipatory deficit in food wanting. Melancholic MDD is associated with HPA hyperactivity (55), and preclinical studies have shown that ghrelin regulates stress-induced anxious behaviour via the HPA axis (139), supporting a link between homeostatic signals and affective states (75) Such mechanisms may contribute to the anti-depressive effects of fasting interventions (140,141). Taken together, capitalising on metabolic signals might provide better treatments for motivational deficits in depression (80,142).

## 5. Limitations

Despite notable strengths, several study limitations should be addressed in future work. First, parsing reward behaviour into different facets revealed differences between cued anticipation and consummation, but the strong effect of reward proximity on group differences was unexpected. Future research may systematically vary additional components such as proximity and probability or certainty of the reward outcome (28,143,144). Second, we assessed inter-individual differences in fasting levels of hormones and meal-related changes in hormone levels after the consummatory phase could reveal additional contributions to the regulation of reward function. Relatedly, the absence of differences in peripheral hormone levels does not preclude differences in central levels or central sensitivity (69,145). Interventional studies administering insulin or ghrelin will help substantiate the link between metabolism and reward processing in MDD. Third, the taste test consisted of palatable food snacks that might not generalise to other rewards. While we did not find specific associations of reward ratings with single items of the SHAPS, future studies will need to test whether an anticipatory deficit instead of a consummatory deficit pertains to other rewards (e.g., social rewards), as has been shown in schizophrenia (146), potentially using ecological momentary assessments. Fourth, we assumed linear relationships in our sample since it was metabolically healthy. However, non-linear relationships are common in medicine and plausible and larger studies should test for non-linear associations, especially if they include patients with metabolic disorders (e.g., type 2 diabetes). Fifth, given the heterogeneity of MDD symptom profiles (54), our use of an atypical balance score captured well-documented differences among patients. Still, the atypical balance score does not consider the DSM-5 mood reactivity criteria for atypical depression, and heterogeneity of the construct is hindering evidence synthesis (147). Thus, future research could use symptom networks instead (148), which require much larger samples though. Lastly, study participation introduces potential selection bias, as individuals who choose to participate may differ systematically from those who do not, which might affect the generalizability of the findings. Information bias may arise from the self-reported outcomes as participants’ ratings of food liking and wanting could be influenced by various reporting biases. Despite our efforts to control for confounders (i.e., BMI, age, and sex), residual confounding may arise from unmeasured or inadequately measured variables, potentially influencing the observed associations (149).

## 6. Conclusion

Anhedonia is a core symptom of depression, yet it may conflate discernable facets of reward function, calling for distinct mechanistic therapies. Here, we disentangled anticipatory and consummatory phases of reward processing to show that depression and anhedonia are characterised by blunted reward anticipation rather than an inability to derive pleasure from rewards. Crucially, we found that wanting already improves with the proximity of the food reward, pointing to a motivational deficit that is corrected by larger consummatory increases compared to healthy individuals. In line with the motivational role of ghrelin, our results highlight that altered gut-brain signalling may contribute to blunted reward function across phases, which may contribute to the symptoms of melancholic MDD. To conclude, precision-oriented behavioural assessments may pave the way towards optimised treatments of reward deficits to improve the quality of life of patients with anhedonia. Based on our findings, encouraging patients with MDD to deliberately experience rewards by removing potential motivational roadblocks may provide a surprisingly straightforward improvement that can be incorporated into cognitive-behavioural treatment modules.

## Data Availability

All data produced in the present study are available upon reasonable request to the authors.

## Data and code sharing statement

All data (anonymised) for the analysis and the code is available: https://github.com/neuromadlab/TasteTest_depression

## Acknowledgement

We thank Ebru Sarmisak, Anne Schiller, Antonia Schlaich, and Rauda Fahed for help with data acquisition. We also thank Stephanie Ebbinghaus who kindly helped with running the ELISA tests. The study was supported by DFG KR 4555/7-1, KR 4555/9-1, KR 4555/10-1, and & WA 2673/15-1. Figures were created with BioRender.com.

## Contributors

**Corinna Schulz:** Formal Analysis, Visualization, Project administration, Investigation, Writing-Original draft preparation, Reviewing and Editing. **Johannes Klaus**: Project administration, Investigation. **Franziska Peglow**: Investigation. **Anne Kühnel**: Writing-Reviewing and Editing. **Sabine Ellinger**: Investigation, Writing-Reviewing and Editing. **Martin Walter**: Conceptualization, Methodology, Funding acquisition. **Nils B. Kroemer**: Conceptualization, Methodology, Funding acquisition, Supervision, Project administration, Validation, Writing-Reviewing and Editing. All authors read and approved the final version of the manuscript. C.S. and N.B.K. have accessed and verified the data.

