## Supplemental Material for "Blunted anticipation but not consummation of food rewards in depression"

**Supplementary Material**

**SI1. Recruitment flowchart**

From 05.10.2021 until 31.05.2023 (time of data freeze), 697 participants were screened on the telephone for initial eligibility for several studies running in the lab, including the one described here. 292 participants were found to be potentially suitable for this study, of which 133 were invited to participate in the study described here. Of these, 20 participants dropped out before their first session, and 3 dropped out during the study (of which 1 dropped out due to exhaustion). Additionally, 7 participants were excluded during the study (5 because they were identified with past or current severe substance abuse, 1 because of concurrent PTSD, 1 because of breastfeeding). The recruitment process resulted in 103 participants out of the 133 invited, yielding a response recruitment rate of approximately 77%.

20 dropouts before S1

3 dropouts during

7 exclusions during

Initial telephone screening (multiple studies) N = 697

Potentially eligible N = 292

Invited current study N = 133

Final sample N = 103

| **SI2. Participant comorbidities** | | | |
| --- | --- | --- | --- |
| *Participant comorbidities* |  |  |  |
| Characteristic | HCP (N=51) | MDD (N=52) | **Overall (N=103)** |
| **Obsessive compulsive disorder (lifetime)** | 1 (2.0%) | 5 (9.6%) | 6 (5.8%) |
| **Post-traumatic stress disorder (lifetime)** | 1 (2.0%) | 4 (7.7%) | 5 (4.9%) |
| **Attention Deficit Disorder** |  |  |  |
| Inattentive type | 4 (7.8%) | 1 (1.9%) | 5 (4.9%) |
| Combined type | 0 (0%) | 4 (7.7%) | 4 (3.9%) |
| **Binge eating disorder (lifetime)** | 1 (2.0%) | 1 (1.9%) | 2 (1.9%) |
| **Alcohol substance use disorder** |  |  |  |
| Lifetime | 7 (13.7%) | 8 (15.4%) | 15 (14.6%) |
| Severity: moderate   (else light) | 0 (0%) | 2 (3.8%) | 2 (1.9%) |
| Current | 3 (5.9%) | 6 (11.5%) | 9 (8.7%) |
| **Other substance use disorder** |  |  |  |
| Lifetime | 2 (3.9%) | 4 (7.7%) | 6 (5.8%) |
| Severity: moderate   (else light) | 1 (2.0%) | 1 (1.9%) | 2 (1.9%) |
| Current | 2 (3.9%) | 2 (3.8%) | 4 (3.9%) |
| **Social anxiety** | | 11 (21.2%) | 11 (10.7%) |
| **Generalized anxiety** |  | 6 (11.5%) | 6 (5.8%) |
| **Panic disorder** |  | 8 (15.4%) | 8 (7.8%) |
| *Note.* Listed are comorbidities that were no exclusion criteria. Current refers to fulfilment of diagnostic criteria within the last 12 months. Severe substance abuse was excluded. Social anxiety, generalized anxiety, and panic disorder are only mentioned for MDD to strengthen that they were exclusion criteria for HCPs. Values are counts with percentage of respective group. | | | |

**SI3. Atypical Balance Score**

The atypical items of the SIGH-ADS include social withdrawal, weight gain, appetite increase, increased eating, carbohydrate craving or eating, hypersomnia, fatigability, and diurnal variation type B (i.e., mood or energy dips in the afternoon). Following (Williams & Terman, 2003) we calculated an atypical balance score from the SIGH-ADS as follows: total 8-items Atypical Symptoms score divided by the total 25-item SIGH-ADS score (i.e., 17-item Hamilton score + 8-item Atypical Symptom score), multiplied by 100. Thus, the atypical balance score represents the percentage of atypicality, ranging from 0 (minimum) to 100 % (maximum). In our MDD sample, the scores were approximately normal distributed and ranged from 20 to 60 (mean = 40,21, median = 39,64, sd = 8,28). We stratified the MDD sample into participants with low atypical MDD (below median) versus high atypical MDD (above median), allowing to include the categorial *Atypical Group Factor* (HCP vs. melancholic MDD vs. atypical MDD) to test across the whole sample. We complemented the analysis using the atypical balance score as a continuous measure by setting the scores of HCPs to zero and group-centering the scores before including the atypical balance score as a continuous factor.

**SI4. Log-transformation of hormone values**

Hormone values were log-transformed due to the skewness of raw values (Kroemer et al., 2013). The TyG is already log-transformed (Unger et al., 2014), likewise, we log-transformed the HOMA-IR (Okita et al., 2013). Shapiro-Wilk tests for normality indicated approximately normal distributions for residualised log-transformed values of acyl ghrelin (*W* = .99, *p* = .77), des-acyl ghrelin (*W* = .98, *p* = .09), HOMA-IR (*W* = .99, *p* = .31), insulin (*W* = .99, *p* = .47), and glucose (*W* = .98, *p* = .29), but not for TyG (*W* = .96, *p* = .001). However, visual inspection and QQ-Plots indicate that TyG is approximately normally distributed. Additionally, the Kolmogorov-Smirnov tests indicate normal distribution as well (D = .07, p = .61).

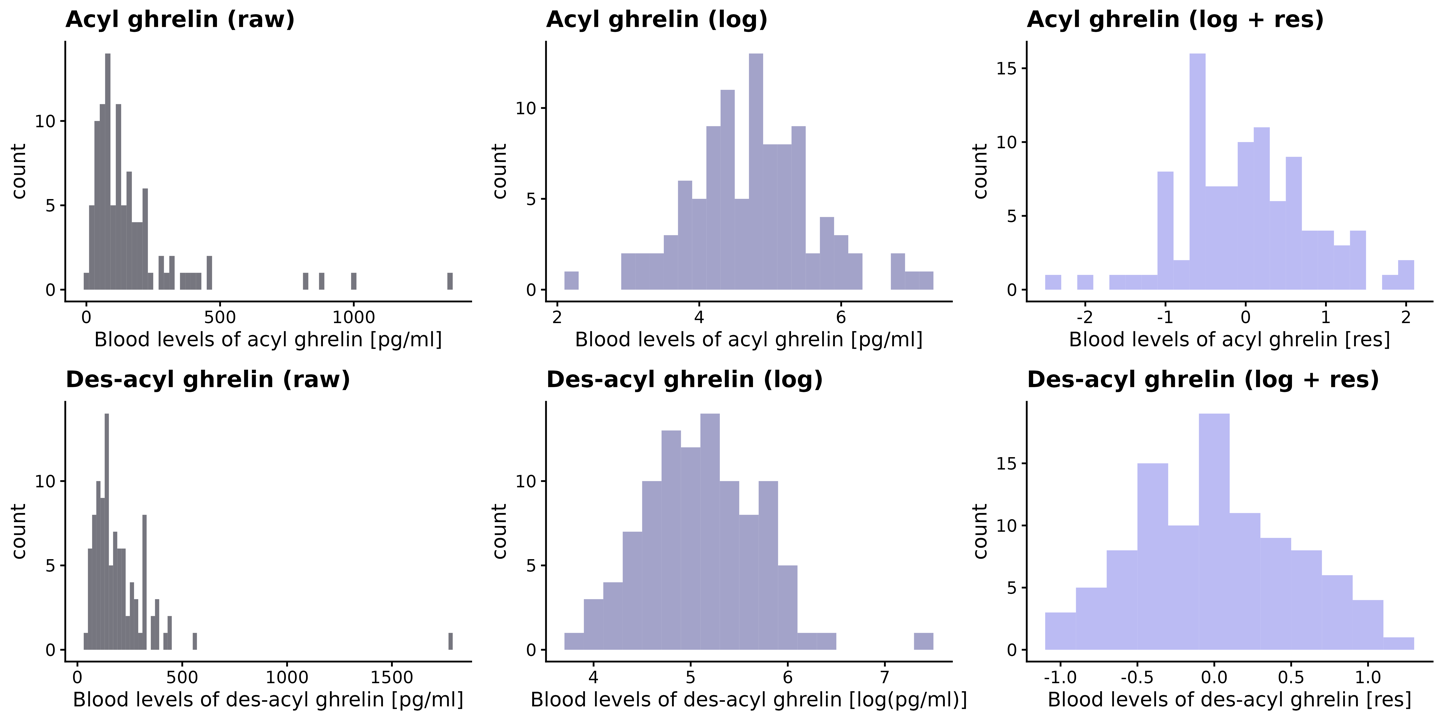

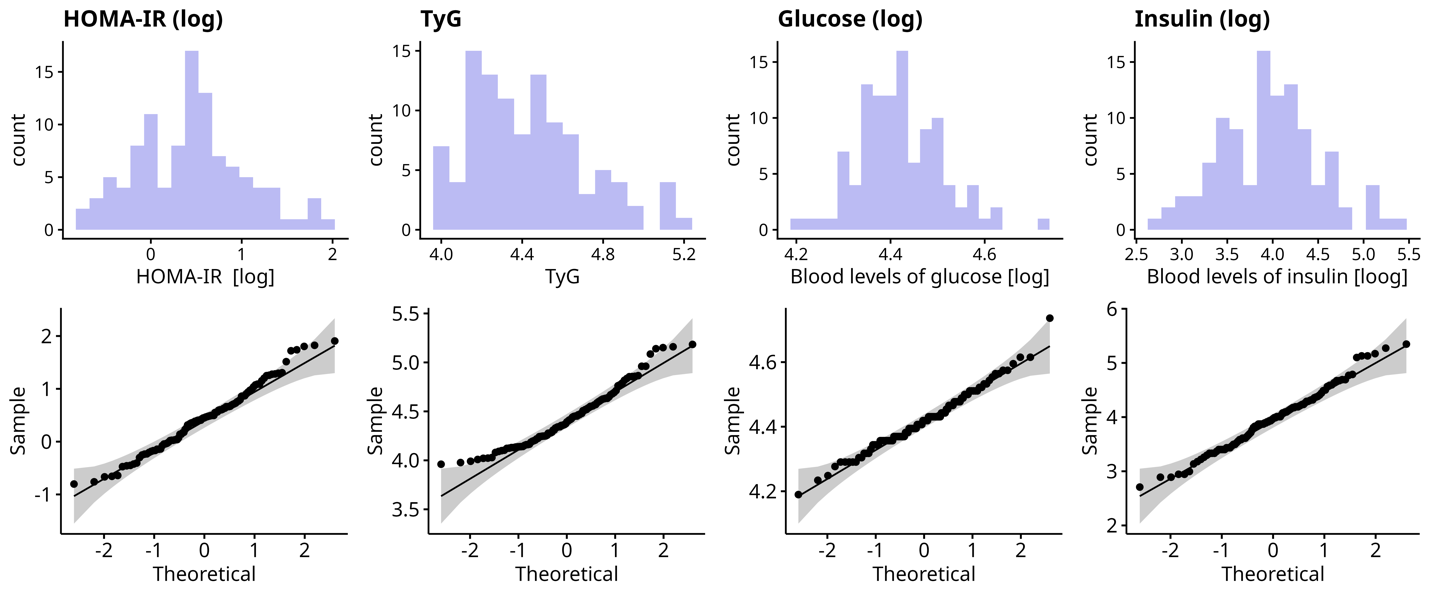

**Figure S3.** Log transformation to achieve approximately normal distributed values. The upper panels show the distribution of acyl- and des-acyl ghrelin before (raw) and after log transformation. Lower panels show approximately normal distribution and corresponding QQ-Plots.

**SI5. Primary and exploratory analyses specifications**

| (1) + (2)   LME Model 1-4:  Rating ~ **Symptom***Phase + Snack + cBMI + cAge + cSex + (1 + Phase + Snack \| ID) | | |
| --- | --- | --- |
| **Rating/ Symptom** | **Depression**  **(MDD vs. HCP)** | **Anhedonia**  **(SHAPS; continuous)** |
| **Liking** | Model 1 | Model 3 |
| **Wanting** | Model 2 | Model 4 |
| (3)  Multivariate Test:  (Liking, Wanting) ~ Horm. Value + MDD*Phase + cBMI + cAge + cSex | | |

**Exploratory analyses:**

Depression subtypes:
Rating ~ **Subtype***Phase + Snack + cBMI + cAge + cSex + (1 + Phase + Snack | ID)
🡪 FDR correction for Main Effects and Interactions each with the Depression Models (Model 1 and Model 2).

Hormonal values:

**Horm. Value** ~ MDD + cSex + cAge + cBMI)
🡪 FDR correction within this family of hypotheses

SHAPS ~ **Horm. Value** + cSex + cAge + cBMI)
🡪 FDR correction within this family of hypotheses

Wanting, Liking, and ghrelin coupling

**Wanting** ~ cLiking * Ghrelin + MDD + cSex + cAge + cBMI

**SI6. Phase codings**

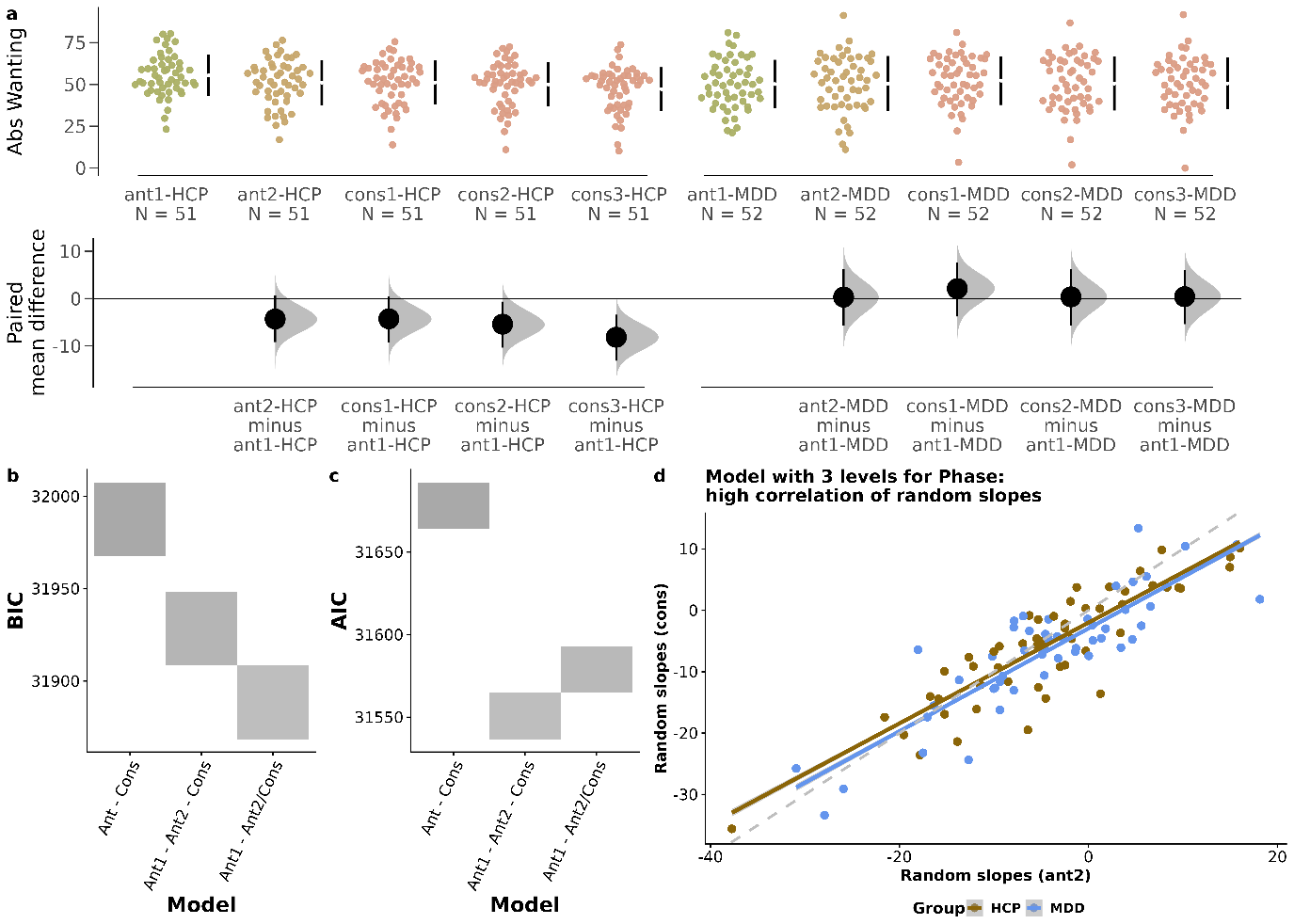

As per hypothesis, we first modelled the phase factor as a 2-level factor with the first two phases (food cues, sight and smell of snacks) as one anticipatory phase, and the last three phases (repeated consummation) as one consummatory phase [Ant-Cons]. However, inspecting raw data we found that individuals adjusted their ratings already markedly after the first phase (Fig. S1a), indicating that ratings in the 2^nd^ anticipation phase already differed qualitatively. Followingly, we decided to model the phase factor as a 3-level factor (1^st^ anticipation, 2^nd^ anticipation, consummation) [Ant1-Ant2-Cons] and as a 2- level phase factor (1^st^ anticipation, 2^nd^ anticipation/consummation) [Ant1-Ant2/Cons] and compared the models [Fig. S1b-d]. First, we found that the latter two models performed better than the original model in a model comparison using the Bayesian and Akaike Information criteria (Fig. S1b-c), indicating a better fit-complexity trade-off. Second, we found that the random slopes correlation for the 3-level phase factor was very high (*r* = 0.83; Fig. S1d), suggesting that the additional third phase does not differ qualitatively. Therefore, we used the 2-level phase factor to separate first anticipation (i.e., cued) from later anticipation (i.e., sight and smell) and consummation for all further analysis. Importantly, the conclusions for the group differences (next paragraph) did not change qualitatively using different phase coding.

**SI7. BDI anhedonia and liking, wanting**

As some items from the BDI tap into anhedonia (Pizzagalli et al., 2005), we also investigated the BDI anhedonia subscore, partially replicating the pattern for SHAPS. We replicated results for reduced initial wanting (*b* = -3.59, [CI: -7.01; -0.11], *p* = .046) but not increases with proximal food (*b_BDI_anhxPhase_* = 2.74, [CI: -0.47; 5.95], *p* = .098). Further replicating results with SHAPS, BDI anhedonia was not associated with liking (*b* = -2.66, [CI: -7.26; 1.95], *p* = .26) or changes in liking with consummation (*b_BDI_anhxPhase_* = 1.39, [CI: -1.78; 4.56], *p* = .39).

**SI8. Accounting for liking in the models for wanting (sensitivity analysis)**

(1) **Wanting** ~ MDD * Phase * c**Liking** + Snack + cBMI + cAge + cSex +

(1 + cLiking + Snack + Phase | ID)

(2) **Wanting** ~ cSHAPS * Phase * c**Liking** + Snack + cBMI + cAge + cSex +

(1 + cLiking + Snack + Phase | ID)

**Including liking in the models for wanting does not alter the main conclusions of lower wanting in MDD and anhedonia and increases in wanting with consummation in MDD and anhedonia, respectively.**

| **model** | **term** | **estimate** | **Std. Error** | **p-value** |
| --- | --- | --- | --- | --- |
| (1) | MDD | **-5.01** | **2.32** | **.03** |
|  | Liking | **0.19** | **0.05** | **.0005** |
|  | MDD * Liking | -0.01 | 0.07 | .85 |
|  | MDD * Phase | **6.12** | **2.16** | **.005** |
|  | Liking * Phase | **0.18** | **0.04** | **<.001** |
|  | Liking * Phase * MDD | -0.11 | 0.06 | .067 |
| **(2)** | SHAPS | **-0.38** | **0.14** | **.008** |
|  | Liking | **0.18** | **0.03** | **<.001** |
|  | SHAPS * Liking | -0.006 | 0.004 | .16 |
|  | SHAPS * Phase | **0.29** | **0.13** | **.035** |
|  | Liking * Phase | **0.12** | **0.03** | **<.001** |
|  | Liking * Phase * SHAPS | 0.004 | 0.004 | **.**33 |

**SI9. Influence of potential violation of heteroscedasticity**

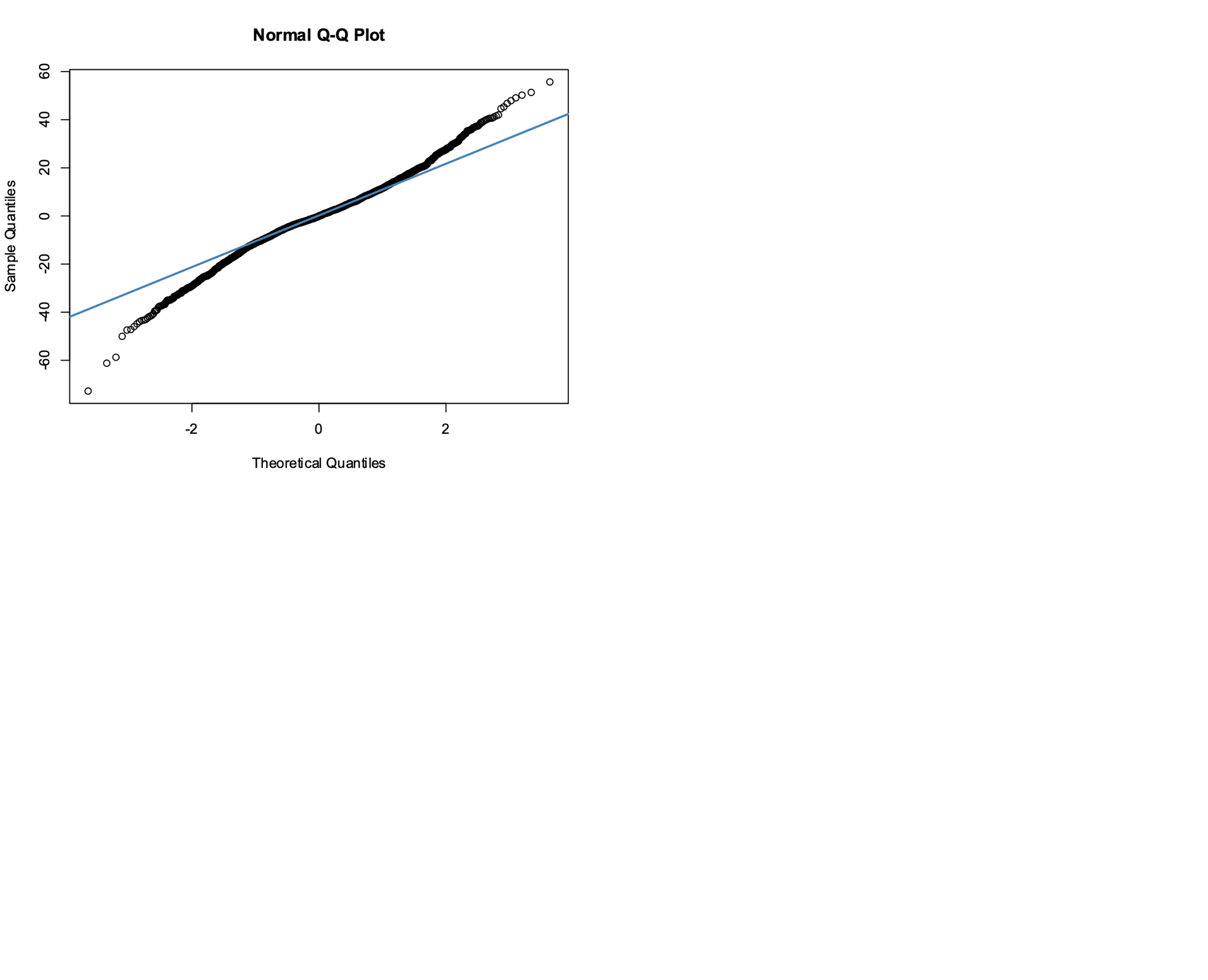

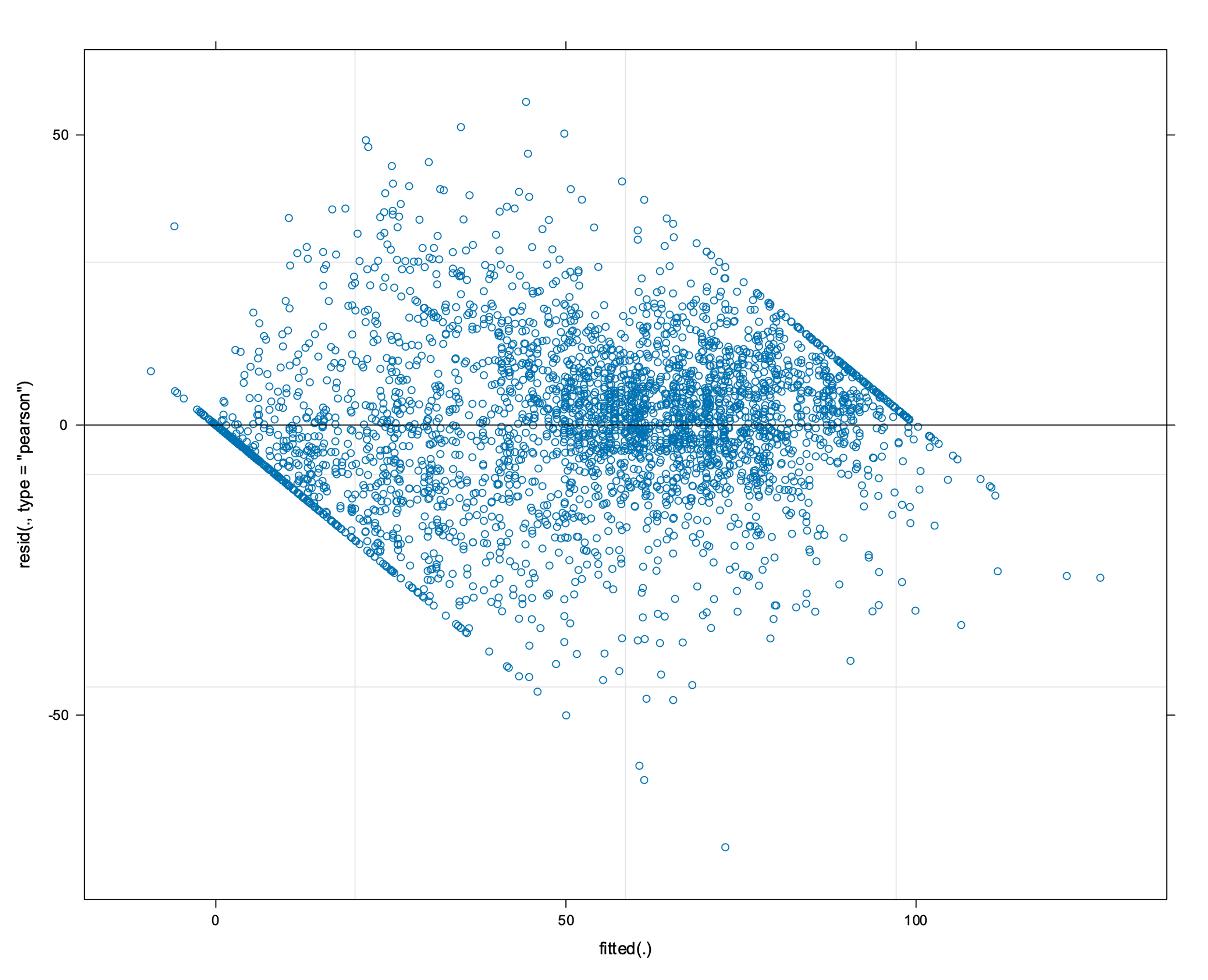
**SI8**

**Figure S8.** The left panel shows the fitted vs. predicted residual plot exemplary for the liking depression model, indicating some heteroscedasticity. The right panel shows the QQ Plot for the same model, confirming approximate normality of residuals.

**Table S9.**

| **model** | **term** | **test** | **estimate** | **se** | **lower** | **upper** |
| --- | --- | --- | --- | --- | --- | --- |
| **Liking and depression** | MDD | P | 0.21 | 3.44 | -6.54 | 6.96 |
|  |  | NP | 0.21 | 3.29 | -6.32 | 6.56 |
|  |  | **bias** | **0** | **+0.15** |  | |
|  | MDD*  Phase[consummation] | P | 2.65 | 2.33 | -1.91 | 7.23 |
|  |  | NP | 2.66 | 2.32 | -1.75 | 7.34 |
|  |  | **bias** | **+0.01** | **-0.01** |  | |
| **Liking and SHAPS** | SHAPS | P | -0.36 | 0.20 | -0.76 | 0.04 |
|  |  | NP | -0.36 | 0.21 | -0.78 | 0.06 |
|  |  | **bias** | **0** | **+0.01** |  | |
|  | SHAPS*  Phase[consummation] | P | 0.25 | 0.14 | -0.03 | 0.52 |
|  |  | NP | 0.25 | 0.13 | -0.004 | 0.52 |
|  |  | **bias** | **0** | **-0.01** |  | |
| **Wanting and depression** | MDD | P | -5.17 | 2.56 | -10.19 | -0.15 |
|  |  | NP | -5.17 | 2.64 | -10.40 | -0.08 |
|  |  | **bias** | **0** | **-0.08** |  | |
|  | MDD*  Phase[consummation] | P | 6.73 | 2.32 | 2.19 | 11.28 |
|  |  | NP | 6.74 | 2.41 | 2.17 | 11.60 |
|  |  | **bias** | **+0.01** | **-0.09** |  |  |
| **Wanting and SHAPS** | SHAPS | P | -0.40 | 0.15 | -0.70 | -0.10 |
|  |  | NP | -0.40 | 0.17 | -0.73 | -0.08 |
|  |  | **bias** | **0** | **+0.02** |  | |
|  | SHAPS*  Phase[consummation] | P | 0.30 | 0.14 | 0.02 | 0.58 |
|  |  | NP | 0.30 | 0.13 | 0.06 | 0.57 |
|  |  | **bias** | **0** | **-0.01** |  | |

**Table S9.** **Bias of non-parametric method using wild bootstrapped results for linear mixed effects models (N*_B_* = 1000).** Wild bootstrapping makes no distribution assumptions and allows for heteroskedasticity (Magnudno and Giannerini, 2015). For the four models, we show the terms of interest (i.e., fixed effects for MDD or SHAPS and their interaction with phase), the coefficient (estimate), the standard error (se) and the 95% confidence interval (upper, lower) for the parametric test (P) and the non-parametric (NP) bootstrapping. The bias of the parametric method was determined by calculating the difference to the non-parametric parameter estimate and standard error.

**Table SI10. Model results for liking and wanting with depression**

|  | **Liking depression** | | | **Wanting depression** | | |
| --- | --- | --- | --- | --- | --- | --- |
| *Predictors* | *Estimates* | *std. Error* | *p* | *Estimates* | *std. Error* | *p* |
| (Intercept) | 12.65 | 2.45 | **<0.001** | 55.36 | 1.84 | **<0.001** |
| fMDD [MDD] | 0.21 | 3.44 | 0.952 | -5.17 | 2.56 | **0.046** |
| fPhase dicho FCR TT [taste_test] | 1.20 | 1.66 | 0.471 | -5.74 | 1.69 | **0.001** |
| fSnack1 | 0.56 | 1.70 | 0.741 | 0.10 | 1.74 | 0.954 |
| fSnack2 | 26.25 | 2.45 | **<0.001** | 22.96 | 2.19 | **<0.001** |
| fSnack3 | -7.09 | 2.91 | **0.016** | -6.54 | 2.40 | **0.008** |
| fSnack4 | 8.22 | 2.74 | **0.003** | 9.27 | 2.29 | **<0.001** |
| fSnack5 | 9.51 | 1.52 | **<0.001** | 8.99 | 1.71 | **<0.001** |
| fSnack6 | -17.43 | 3.42 | **<0.001** | -14.19 | 2.58 | **<0.001** |
| cBMI | -0.52 | 0.42 | 0.223 | -0.59 | 0.37 | 0.115 |
| cAge | -0.04 | 0.19 | 0.843 | -0.30 | 0.16 | 0.067 |
| cSex | -3.71 | 2.58 | 0.154 | -3.71 | 2.27 | 0.106 |
| fMDD [MDD] × fPhase dicho FCR TT [taste_test] | 2.66 | 2.33 | 0.257 | 6.74 | 2.32 | **0.004** |
| **Random Effects** | | | | | | |
| σ^2^ | 221.73 | | | 214.74 | | |
| τ_00_ | 276.79 _ID_ | | | 147.01 _ID_ | | |
| τ_11_ | 260.04 _ID.fSnack1_ | | | 273.77 _ID.fSnack1_ | | |
|  | 581.41 _ID.fSnack2_ | | | 455.06 _ID.fSnack2_ | | |
|  | 831.97 _ID.fSnack3_ | | | 556.49 _ID.fSnack3_ | | |
|  | 733.81 _ID.fSnack4_ | | | 503.93 _ID.fSnack4_ | | |
|  | 198.62 _ID.fSnack5_ | | | 263.82 _ID.fSnack5_ | | |
|  | 1166.24 _ID.fSnack6_ | | | 646.77 _ID.fSnack6_ | | |
|  | 102.99 _ID.fPhase_dicho_FCR_TTtaste_test_ | | | 113.24 _ID.fPhase_dicho_FCR_TTtaste_test_ | | |
| ρ_01_ | 0.08 | | | 0.03 | | |
|  | -0.15 | | | -0.03 | | |
|  | 0.05 | | | 0.21 | | |
|  | -0.09 | | | -0.03 | | |
|  | 0.06 | | | 0.01 | | |
|  | 0.06 | | | -0.03 | | |
|  | -0.62 | | | -0.37 | | |
| ICC | 0.79 | | | 0.73 | | |
| N | 103 _ID_ | | | 103 _ID_ | | |
| Observations | 3605 | | | 3605 | | |
| Marginal R^2^ / Conditional R^2^ | 0.187 / 0.826 | | | 0.207 / 0.789 | | |

**SI11. Comparison 2- versus 3-level phase codings**

| **Phase coding** | **Model** | **Term** | **Estimate** | **Std. error** | **p-value** |
| --- | --- | --- | --- | --- | --- |
| **3 phases** | **Wanting** | MDD | **-5.22** | **2.56** | **.045** |
|  |  | MDD*Phase[anticipation2] | 4.66 | 2.63 | .079 |
|  |  | MDD*Phase[consumption] | **7.43** | **2.36** | **.002** |
|  |  | SHAPS | **-0.40** | **0.15** | **.0097** |
|  |  | SHAPS*Phase[anticipation2] | 0.22 | 0.16 | .17 |
|  |  | SHAPS*Phase[consumption] | **0.25** | **0.009** | **.027** |
|  | **Liking** | MDD | 0.21 | 3.44 | .95 |
|  |  | MDD*Phase[anticipation2] | 1.77 | 2.49 | .48 |
|  |  | MDD*Phase[consumption] | 2.96 | 2.40 | **.**22 |
|  |  | SHAPS | -0.36 | 0.20 | .081 |
|  |  | SHAPS*Phase[anticipation2] | 0.15 | 0.15 | .32 |
|  |  | SHAPS*Phase[consumption] | **0.28** | **0.14** | **.054** |

**Table S11. Similar results for wanting using different codings of the factor phase.** Results are shown for using three phases (ant1 – ant2 – consummation). Results are highlighted in green when they correspond to the results reported using the winning model (Ant1 – Ant2/consummation; reported main manuscript). The table shows that the conclusions derived in the manuscript do not depend critically on choosing the Ant1 – Ant2/consummation model over the (ant 1 – ant2 – consummation).

**SI12. Lower wanting in melancholic MDD**

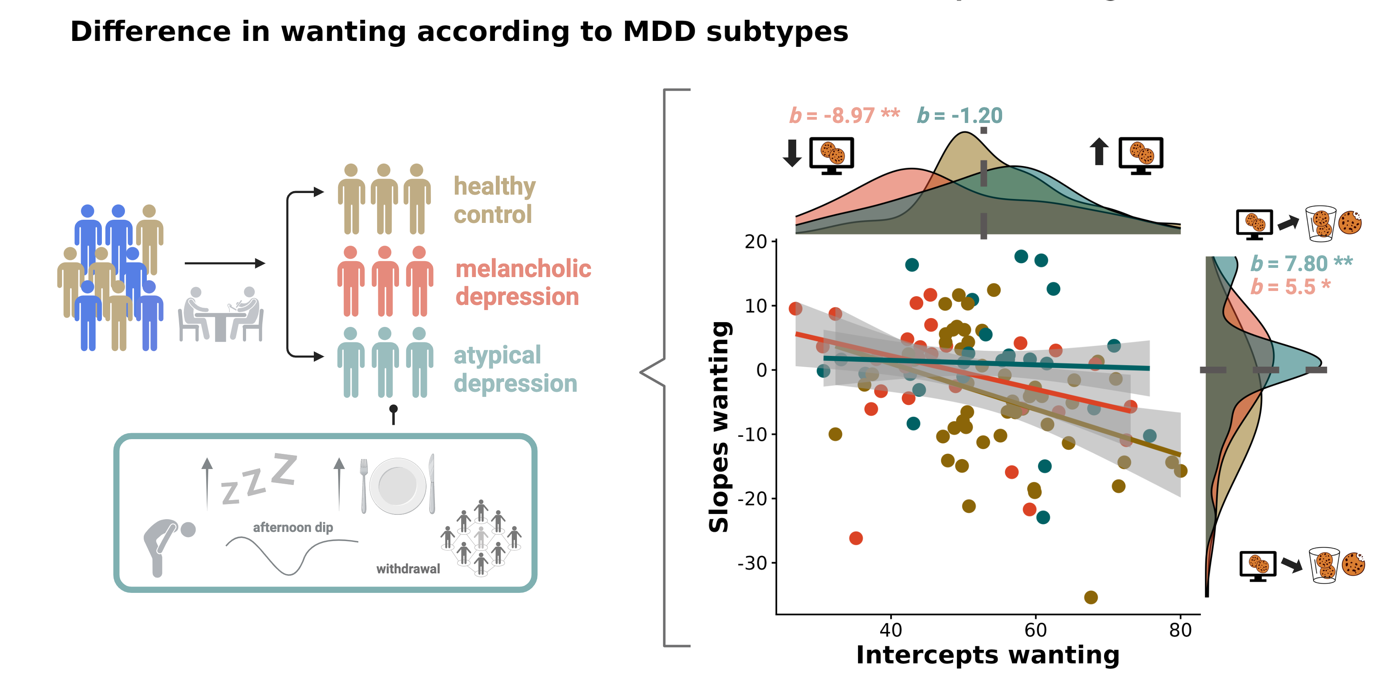

**Fig. S12. Lower wanting in melancholic MDD**. For participants with MDD extend of atypical symptoms were evaluated using the atypical balance score from the SIGH-ADS. Lower wanting during cued anticipation was driven by participants with melancholic (b = -8.97, p = .004) and not atypical MDD (b = -1.20, p = .70). Both, melancholic (b = 5.73, p = .044) and atypical MDD (b = 7.80, p = .008) increased their wanting ratings after cued anticipation.

**Table SI13. Model results for liking and wanting with anhedonia**

|  | **Liking anhedonia** | | | **Wanting anhedonia** | | |
| --- | --- | --- | --- | --- | --- | --- |
| *Predictors* | *Estimates* | *std. Error* | *p* | *Estimates* | *std. Error* | *p* |
| (Intercept) | 12.73 | 1.71 | **<0.001** | 52.74 | 1.29 | **<0.001** |
| cSHAPS sum | -0.36 | 0.20 | 0.081 | -0.40 | 0.15 | **0.010** |
| fPhase dicho FCR TT [taste_test] | 2.55 | 1.17 | **0.032** | -2.34 | 1.23 | 0.060 |
| fSnack1 | 0.56 | 1.70 | 0.741 | 0.10 | 1.74 | 0.954 |
| fSnack2 | 26.25 | 2.45 | **<0.001** | 22.96 | 2.19 | **<0.001** |
| fSnack3 | -7.09 | 2.91 | **0.016** | -6.54 | 2.40 | **0.008** |
| fSnack4 | 8.22 | 2.74 | **0.003** | 9.27 | 2.29 | **<0.001** |
| fSnack5 | 9.51 | 1.52 | **<0.001** | 8.99 | 1.71 | **<0.001** |
| fSnack6 | -17.43 | 3.42 | **<0.001** | -14.19 | 2.58 | **<0.001** |
| cBMI | -0.45 | 0.42 | 0.293 | -0.54 | 0.37 | 0.147 |
| cAge | -0.10 | 0.19 | 0.608 | -0.33 | 0.16 | **0.047** |
| cSex | -3.35 | 2.58 | 0.198 | -3.80 | 2.23 | 0.092 |
| cSHAPS sum × fPhase dicho FCR TT [taste_test] | 0.25 | 0.14 | 0.080 | 0.30 | 0.14 | **0.037** |
| **Random Effects** | | | | | | |
| σ^2^ | 221.72 | | | 214.74 | | |
| τ_00_ | 267.46 _ID_ | | | 140.42 _ID_ | | |
| τ_11_ | 260.05 _ID.fSnack1_ | | | 273.76 _ID.fSnack1_ | | |
|  | 581.62 _ID.fSnack2_ | | | 455.03 _ID.fSnack2_ | | |
|  | 831.83 _ID.fSnack3_ | | | 556.48 _ID.fSnack3_ | | |
|  | 734.28 _ID.fSnack4_ | | | 503.95 _ID.fSnack4_ | | |
|  | 198.54 _ID.fSnack5_ | | | 263.83 _ID.fSnack5_ | | |
|  | 1165.72 _ID.fSnack6_ | | | 646.70 _ID.fSnack6_ | | |
|  | 101.19 _ID.fPhase_dicho_FCR_TTtaste_test_ | | | 117.75 _ID.fPhase_dicho_FCR_TTtaste_test_ | | |
| ρ_01_ | 0.06 | | | 0.04 | | |
|  | -0.14 | | | -0.01 | | |
|  | 0.04 | | | 0.19 | | |
|  | -0.09 | | | -0.03 | | |
|  | 0.10 | | | -0.01 | | |
|  | 0.07 | | | -0.03 | | |
|  | -0.60 | | | -0.37 | | |
| ICC | 0.79 | | | 0.73 | | |
| N | 103 _ID_ | | | 103 _ID_ | | |
| Observations | 3605 | | | 3605 | | |
| Marginal R^2^ / Conditional R^2^ | 0.187 / 0.825 | | | 0.208 / 0.789 | | |

**SI14. SHAPS and atypical depression**

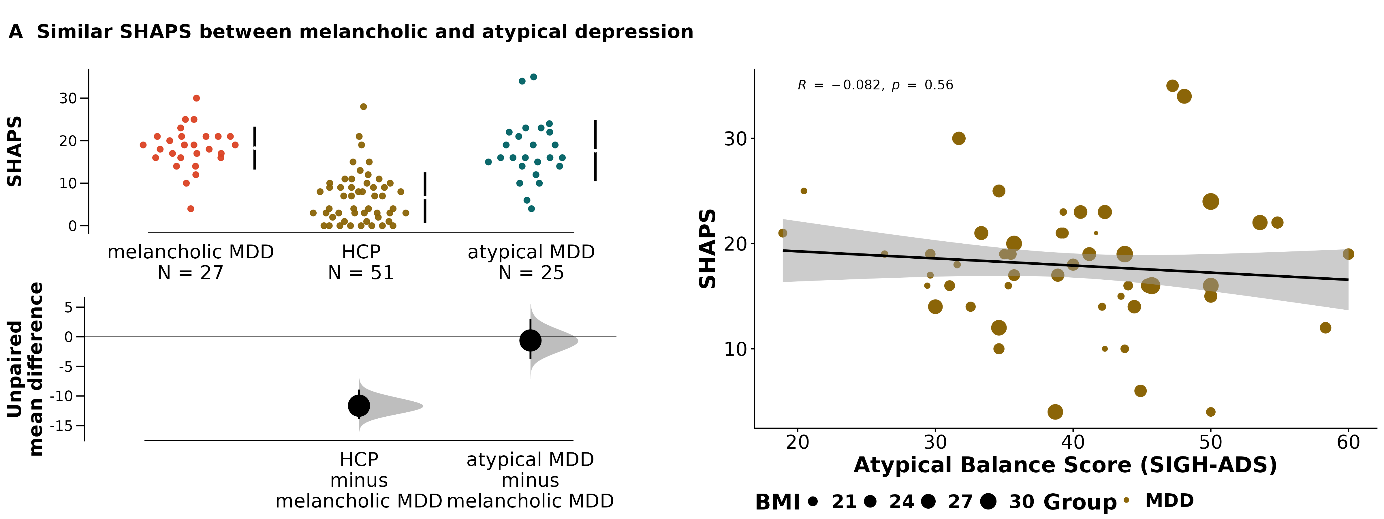

Fig. S4. No differences in SHAPS for depression subtype. Cumming estimation plots show no difference in SHAPS ratings between melancholic and atypical MDD (left). Effect size and bootstrapped 95% Cis are plotted below the raw data. Within participants with depression, atypical balance score was also not associated with SHAPS (*r* = -.082, *p* = .56, right).

**SI15. Neither depression nor anhedonia is characterized by differences in perceived taste**

To evaluate if group differences in wanting were accounted for by differences in perceived taste, we also tested for differences in ratings of intensity, sweetness, saltiness, and savoriness acquired during the consummatory phase of the taste test. Patients with MDD did not differ in their ratings of intensity (*b* = 3.12, *p* = .11), sweetness (*b* = .78, *p* = .43), saltiness (*b* = -1.14, *p* = .27), or savoriness (*b* = -1.35, *p* = .46) compared with HCPs. Likewise, there were no associations with SHAPS or the atypical balance score, except for higher intensity ratings in atypical MDD (*b* = 5.07, *p* = .035) compared to HCPs. Further, intensity ratings did not contribute to the observed differences in wanting between HCPs and depression subtypes (*b_Intensity_* = .0003, *p* = .95, *p*_Intensity x MDD_ = .88) or the absence thereof in liking (*b_Intensity_* = .0005, *p_Intensity_* = .95). These results further corroborate that depression and anhedonia are not associated with altered taste perception per se. Further, as we reported previously in Fahed et al. (2023), patients with MDD did not differ from HCPs in subjective ratings of metabolic state (i.e., hunger, fullness).

**SI16. Depression as anticipatory but not consummatory deficit**

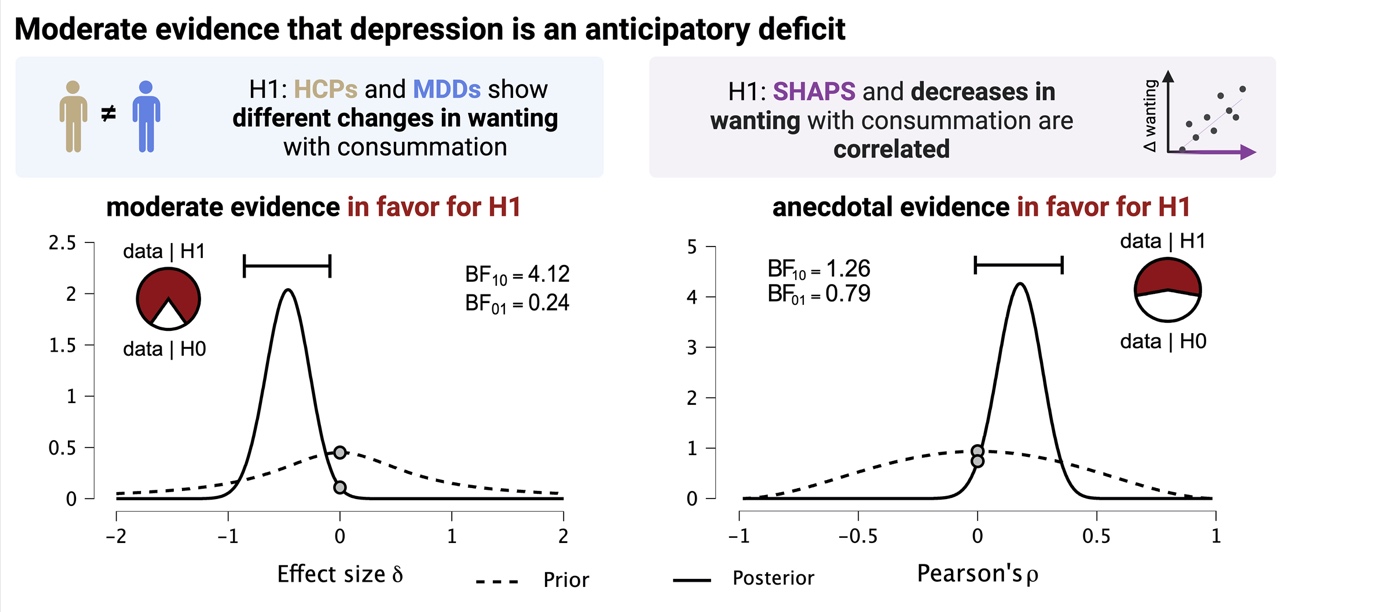

**SI17. Bayes factor robustness check for Independent Samples T-Test**

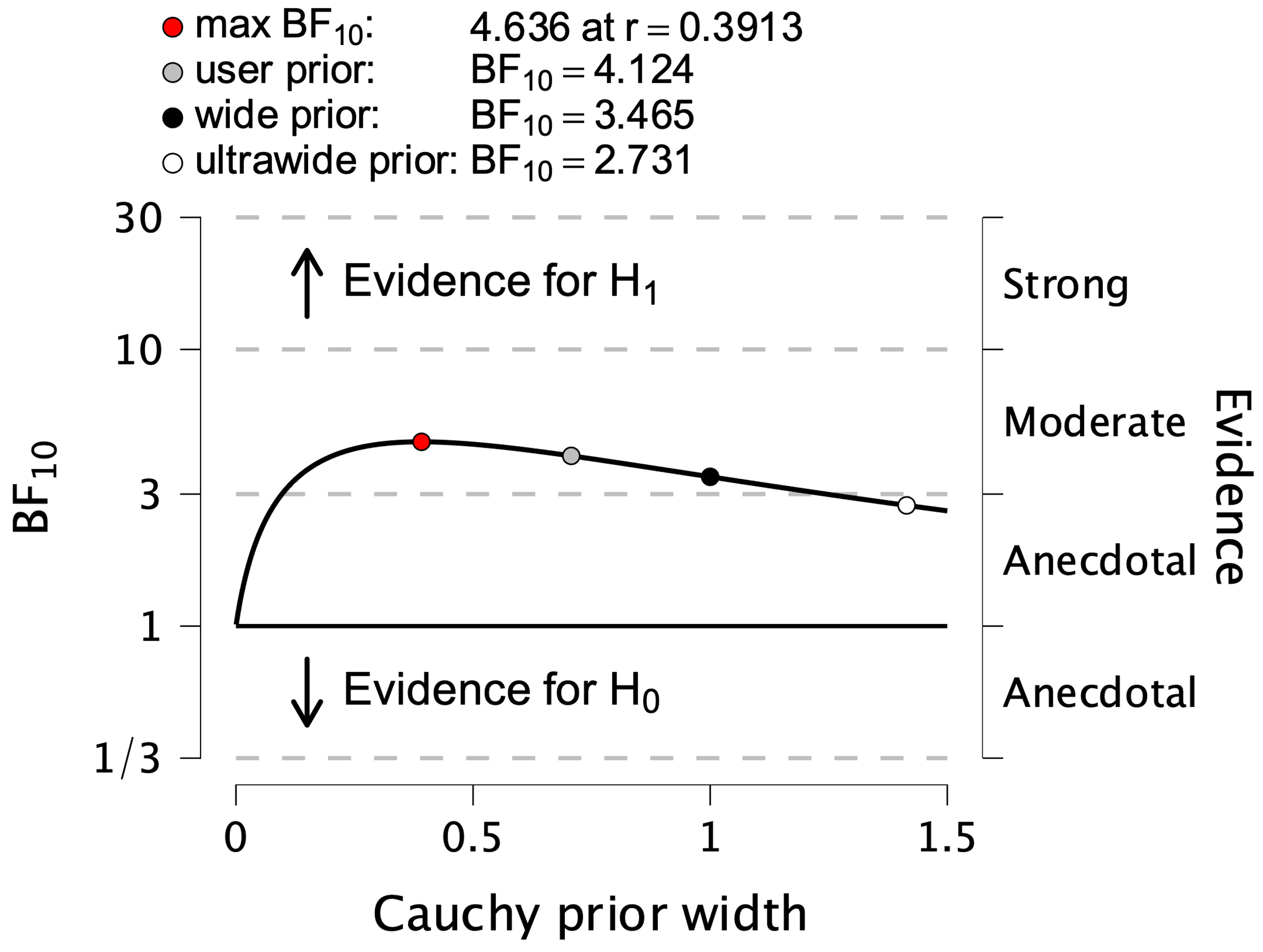

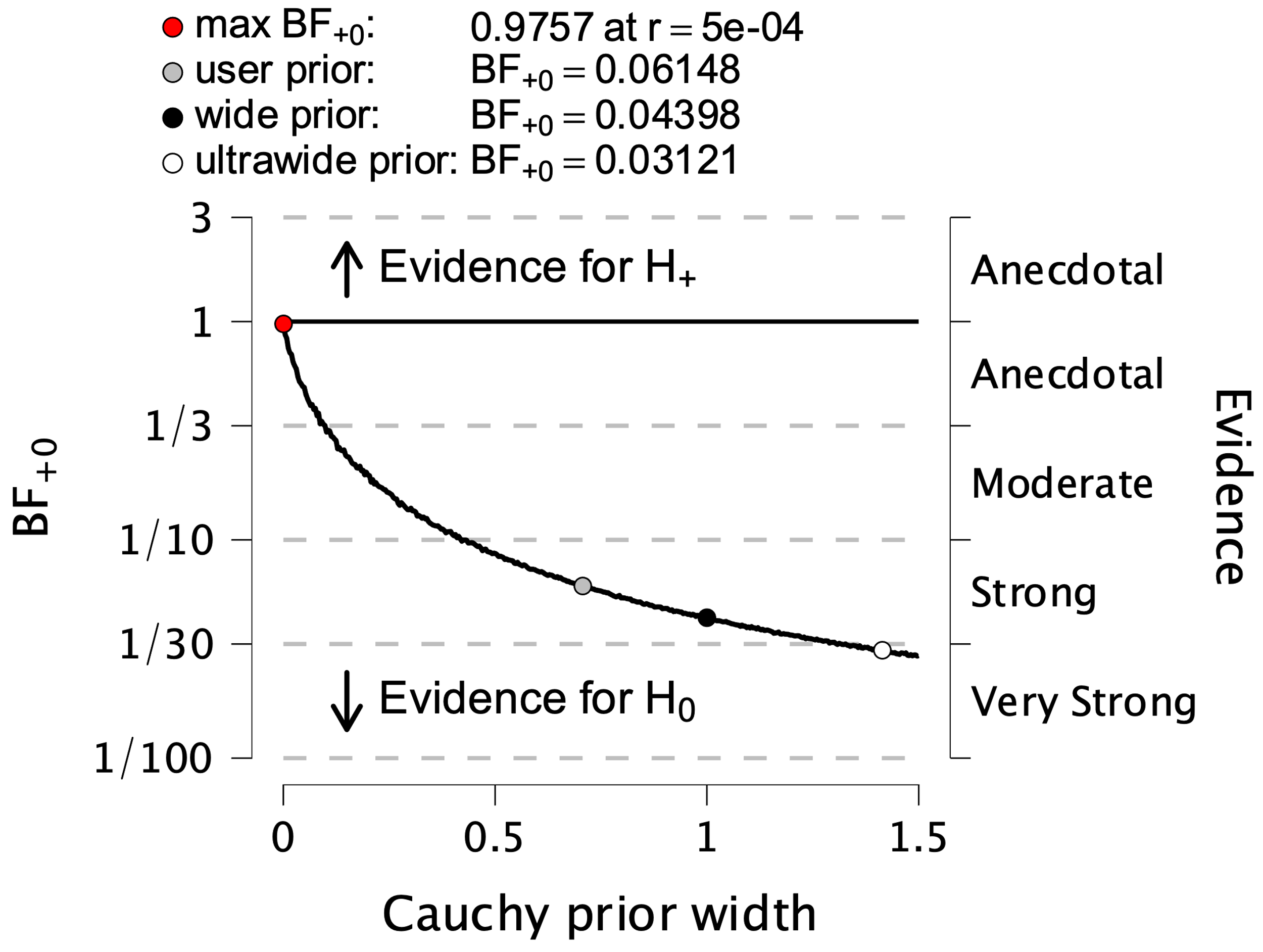

Fig. S17. Robustness Check for the Bayesian Indepent Samples T-Test. Left: Robustness check for the one-sided test that HCPs show greater wanting during consummation than patients with MDD compared to anticipation. Right: Robustness check for the two-sided test that HCPs and patients with MDD differ in their wanting adjustments during consummation.

**Fig. S18 Bayes factor robustness check for Correlation**

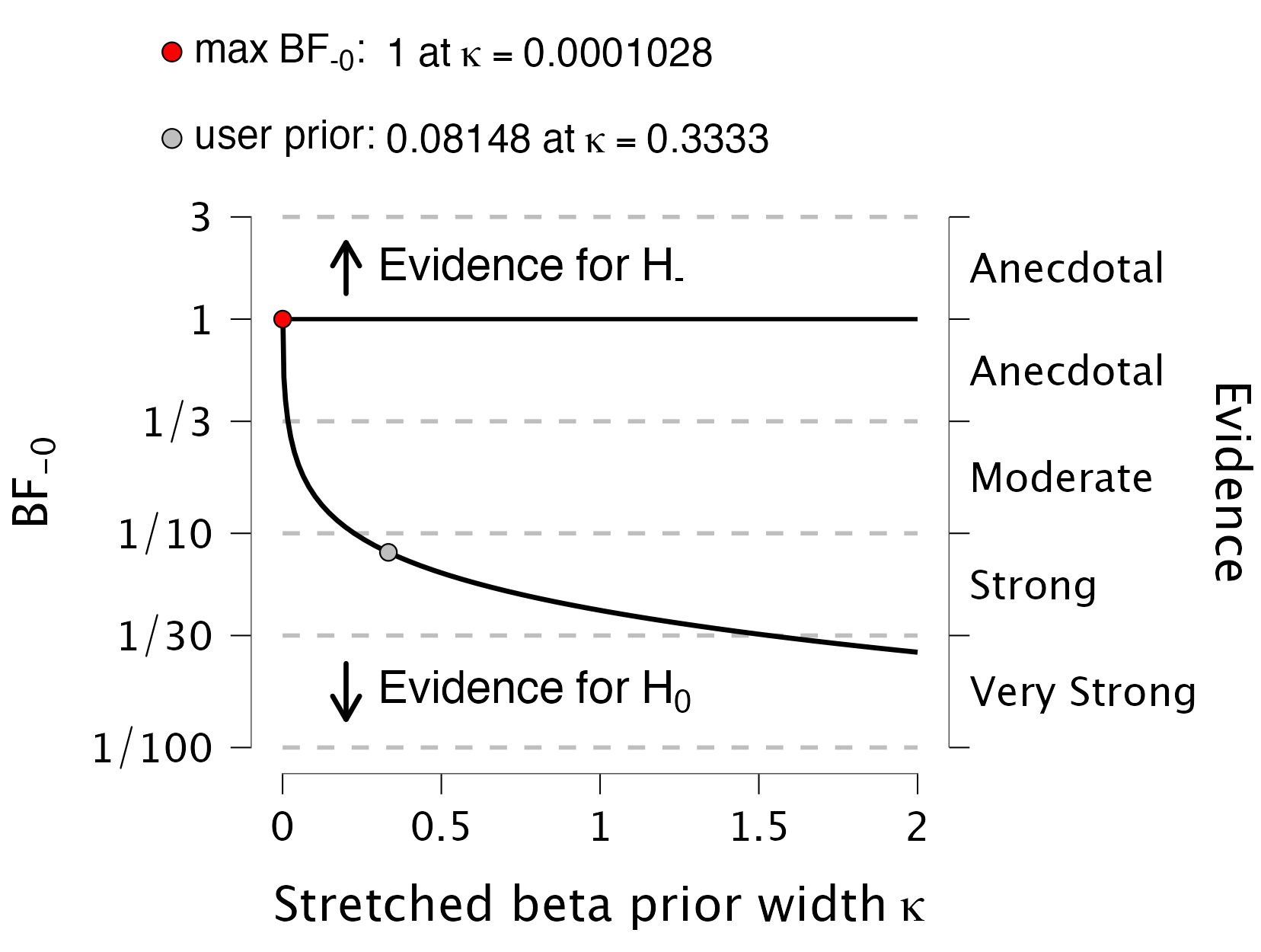

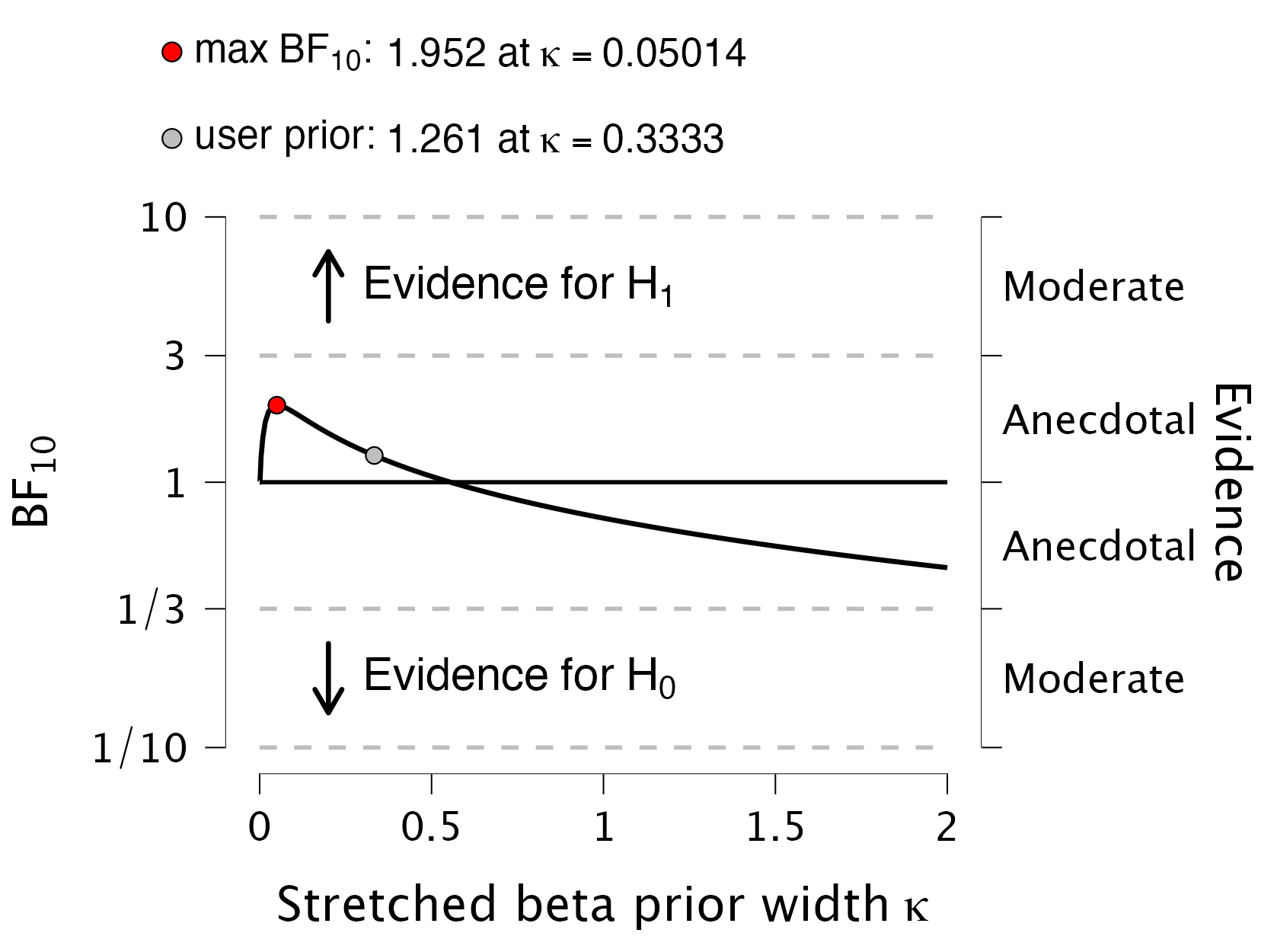

Fig. S18. Robustness Check for the Bayesian Correlation. Left: Robustness check for the directed correlation that SHAPS is negatively associated with changes in wanting with consummation. Right: Robustness check for a correlation test whether SHAPS is correlated with changes in wanting with consummation.

**SI19. Ghrelin and atypical depression**

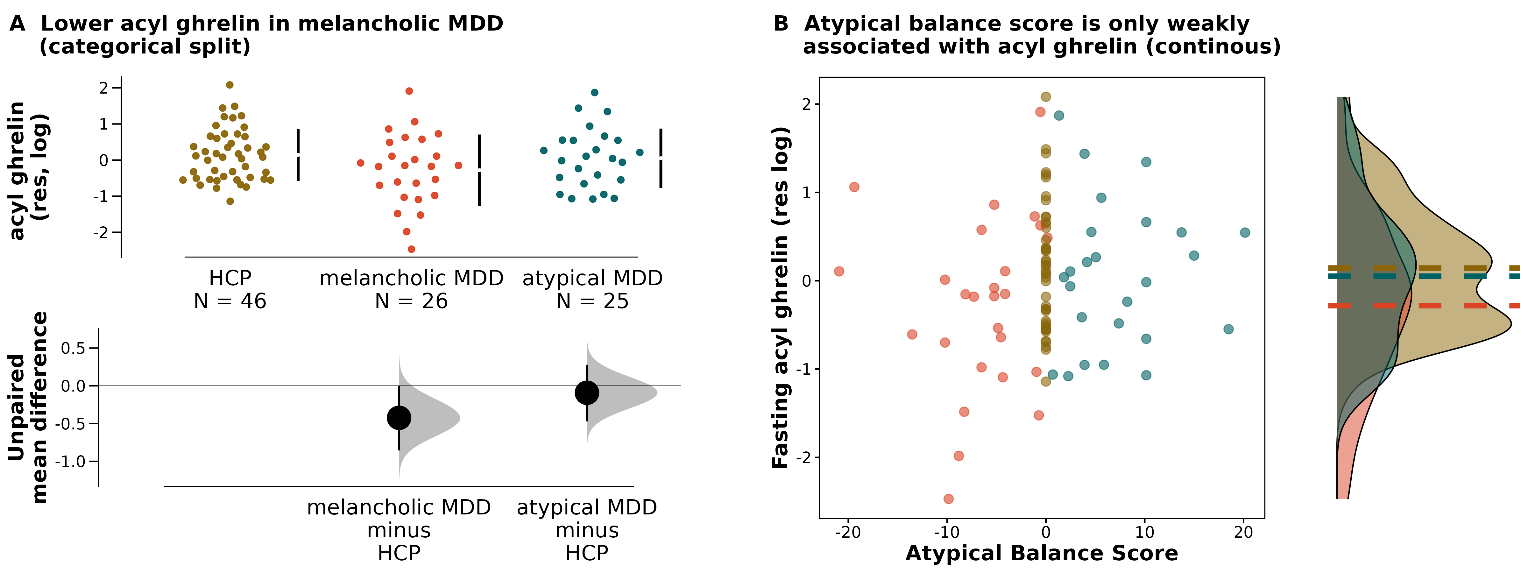

Fig. S8. Cumming estimation plots show lower ghrelin in melancholic but not atypical MDD vs. HCPs (*b* = -.44, *p* = .039, left). Effect size and bootstrapped 95% Cis are plotted below the raw data. The atypical balance score (with HCPs set to zero) was not significantly correlated with ghrelin levels (*r* = .14*, p* = .17*,* right).

**SI20. Ghrelin and SHAPS**

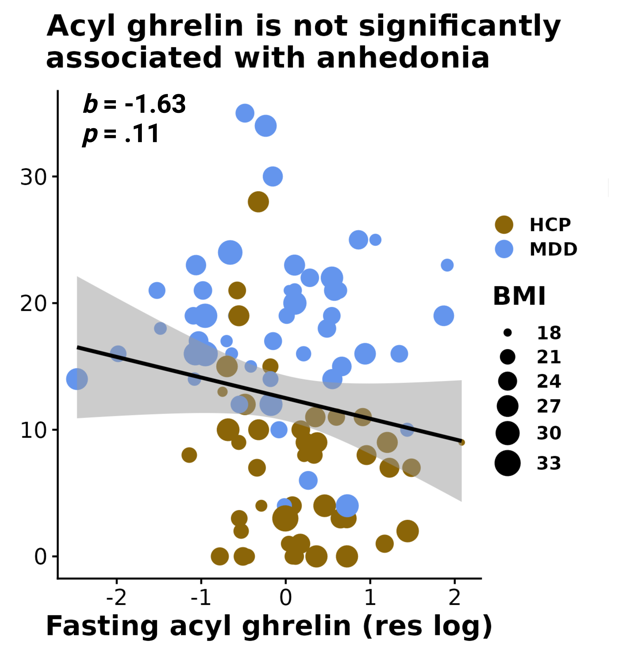

Fig. S9. Acyl ghrelin was only weakly associated with SHAPS ratings.

**SI21. HOMA-IR and SHAPS depending on depression subtype**

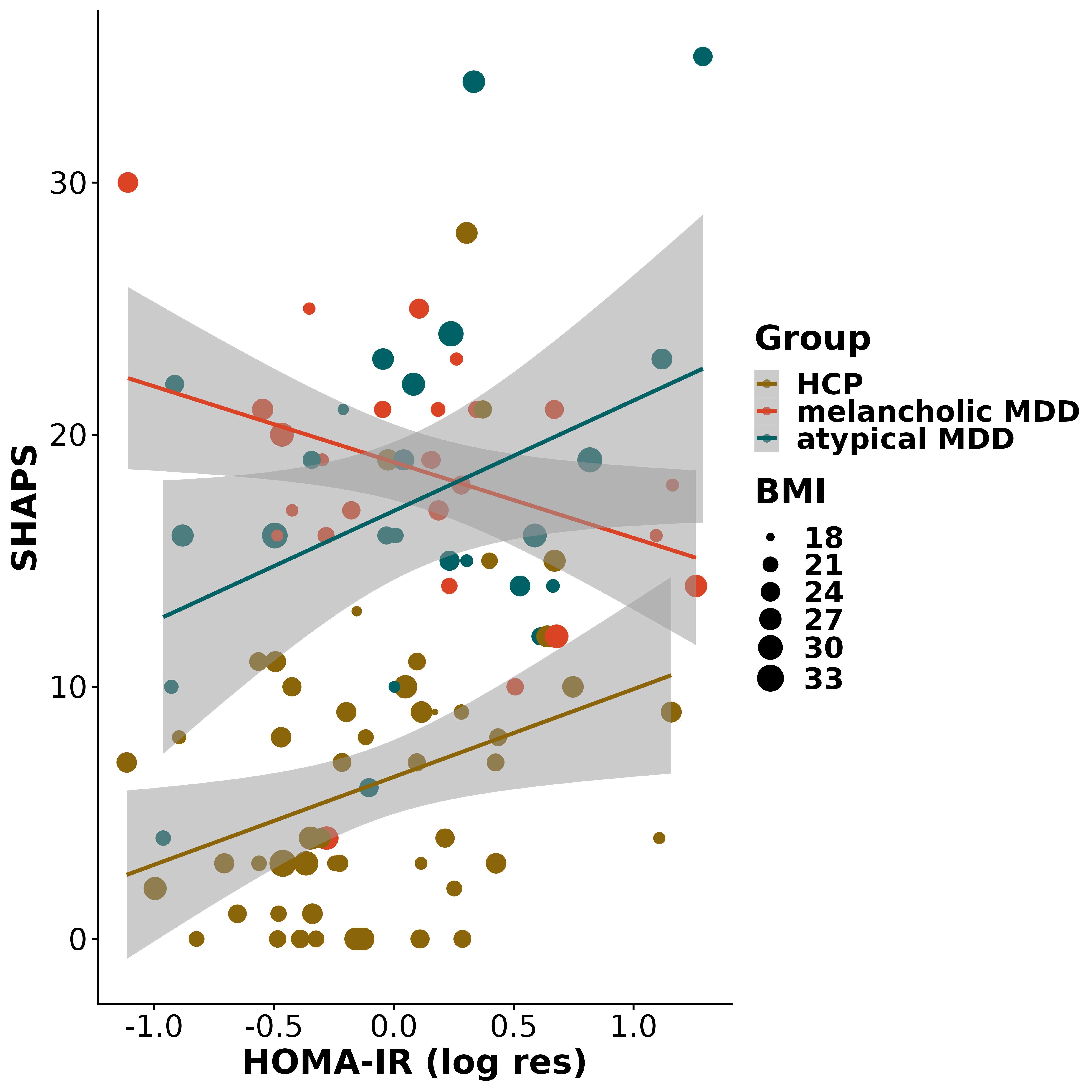

**SI22. Liking and wanting with ghrelin**

Using linear mixed-effects models, we found no main effect of ghrelin on wanting (*b* = 3.11, [CI: -0.05; 6.16], *p* = .051), but on liking (*b* = 4.26, [CI: 0.11; 8.41], *p* = .047), Phase dummy coded). The interaction of ghrelin with phase was not significant for wanting (*b* = 0.39, [CI: -2.57; 3.32], *p* = .80) or liking (*b* = -1.51, [CI: -4.14; 1.40], *p* = .31).

|  | **Liking Ghrelin** | | | **Wanting Ghrelin** | | |
| --- | --- | --- | --- | --- | --- | --- |
| *Predictors* | *Estimates* | *std. Error* | *p* | *Estimates* | *std. Error* | *p* |
| (Intercept) | 12.19 | 1.98 | **<0.001** | 52.30 | 1.65 | **<0.001** |
| fMDD [MDD] | 3.99 | 2.55 | 0.121 | -1.33 | 2.28 | 0.562 |
| res logF AG | 3.88 | 1.70 | **0.025** | 2.87 | 1.39 | **0.042** |
| fPhase dicho FCR TT1 | -1.30 | 0.62 | **0.039** | 1.24 | 0.66 | 0.069 |
| fSnack1 | 0.39 | 1.78 | 0.825 | -0.03 | 1.75 | 0.986 |
| fSnack2 | 25.67 | 2.51 | **<0.001** | 22.44 | 2.26 | **<0.001** |
| fSnack3 | -8.25 | 2.94 | **0.006** | -7.27 | 2.45 | **0.004** |
| fSnack4 | 9.95 | 2.63 | **<0.001** | 10.27 | 2.31 | **<0.001** |
| fSnack5 | 9.64 | 1.57 | **<0.001** | 8.94 | 1.80 | **<0.001** |
| fSnack6 | -16.94 | 3.56 | **<0.001** | -13.83 | 2.69 | **<0.001** |
| cBMI | -0.58 | 0.40 | 0.144 | -0.61 | 0.38 | 0.112 |
| cSex | -3.84 | 2.63 | 0.149 | -5.87 | 2.31 | **0.014** |
| res logF AG × fPhase dicho FCR TT1 | 0.75 | 0.74 | 0.312 | -0.19 | 0.75 | 0.804 |
| cAge |  |  |  | -0.35 | 0.17 | **0.043** |
| res logF AG × cSex |  |  |  | -2.93 | 2.72 | 0.284 |
| res logF AG × cAge |  |  |  | 0.56 | 0.18 | **0.002** |
| cSex × cAge |  |  |  | -0.42 | 0.34 | 0.218 |
| (res logF AG × cSex) × cAge |  |  |  | 0.27 | 0.35 | 0.446 |
| **Random Effects** | | | | | | |
| σ^2^ | 224.67 | | | 213.94 | | |
| τ_00_ | 194.56 _ID_ | | | 118.36 _ID_ | | |
| τ_11_ | 268.44 _ID.fSnack1_ | | | 260.69 _ID.fSnack1_ | | |
|  | 571.42 _ID.fSnack2_ | | | 459.78 _ID.fSnack2_ | | |
|  | 800.08 _ID.fSnack3_ | | | 547.84 _ID.fSnack3_ | | |
|  | 631.28 _ID.fSnack4_ | | | 482.19 _ID.fSnack4_ | | |
|  | 199.27 _ID.fSnack5_ | | | 276.12 _ID.fSnack5_ | | |
|  | 1191.36 _ID.fSnack6_ | | | 667.32 _ID.fSnack6_ | | |
|  | 27.44 _ID.fPhase_dicho_FCR_TT1_ | | | 32.18 _ID.fPhase_dicho_FCR_TT1_ | | |
| ρ_01_ | 0.11 | | | 0.12 | | |
|  | -0.20 | | | -0.16 | | |
|  | 0.15 | | | 0.22 | | |
|  | -0.17 | | | -0.10 | | |
|  | 0.06 | | | 0.08 | | |
|  | 0.05 | | | -0.02 | | |
|  | 0.39 | | | 0.02 | | |
| ICC | 0.78 | | | 0.73 | | |
| N | 97 _ID_ | | | 97 _ID_ | | |
| Observations | 3395 | | | 3395 | | |
| Marginal R^2^ / Conditional R^2^ | 0.196 / 0.823 | | | 0.226 / 0.792 | | |

**SI23. Age, sex, and ghrelin**

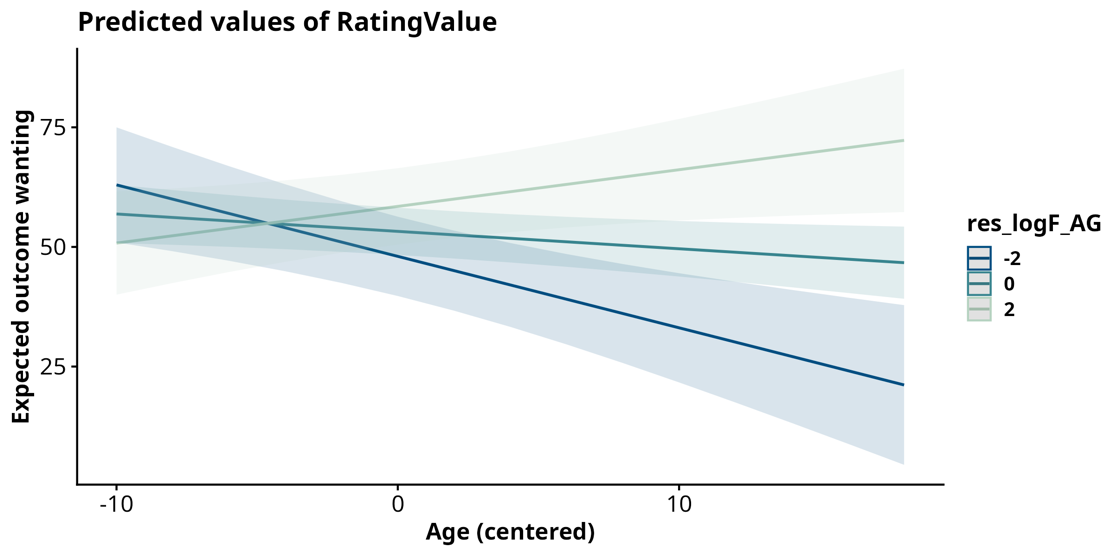

Fig S23. Testing a 3-way interaction of ghrelin, age, and sex showed a significant interaction of ghrelin with age (*b* = 0.56, [CI: 0.21; 0.90], *p*= .002) but not sex (*b* = -2.93, [CI: -8.25; 2.39], *p* = .28; not shown), *uncorrected*. While older participants show lower wanting, this is attenuated with higher ghrelin.

**SI24. HOMA-IR is not associated with wanting or liking**

HOMA-IR was not associated with ratings overall (Pillai’s Trace *V* = .010, *F*(2, 185) = 0.92, *p* = .40).
Separate linear mixed-effect models taking into account the hierarchical structure of the data corroborate this null finding.

|  | **Liking HOMA-IR** | | | **Wanting HOMA-IR** | | |
| --- | --- | --- | --- | --- | --- | --- |
| *Predictors* | *Estimates* | *std. Error* | *p* | *Estimates* | *std. Error* | *p* |
| (Intercept) | 14.02 | 1.42 | **<0.001** | 51.91 | 1.62 | **<0.001** |
| fPhase dicho FCR TT1 | -1.28 | 0.59 | **0.031** | 1.16 | 0.61 | 0.063 |
| res logHOMA | -0.92 | 2.54 | 0.718 | -2.33 | 2.05 | 0.259 |
| fSnack1 | 0.56 | 1.70 | 0.741 | 0.10 | 1.74 | 0.954 |
| fSnack2 | 26.25 | 2.45 | **<0.001** | 22.96 | 2.19 | **<0.001** |
| fSnack3 | -7.09 | 2.91 | **0.016** | -6.54 | 2.40 | **0.008** |
| fSnack4 | 8.22 | 2.74 | **0.003** | 9.27 | 2.29 | **<0.001** |
| fSnack5 | 9.51 | 1.52 | **<0.001** | 8.99 | 1.71 | **<0.001** |
| fSnack6 | -17.43 | 3.42 | **<0.001** | -14.19 | 2.58 | **<0.001** |
| cBMI | -0.49 | 0.42 | 0.252 | -0.59 | 0.37 | 0.116 |
| cAge | -0.07 | 0.19 | 0.726 | -0.30 | 0.16 | 0.074 |
| cSex | -3.38 | 2.58 | 0.193 | -3.64 | 2.27 | 0.112 |
| fPhase dicho FCR TT1 × res logHOMA | 1.24 | 1.07 | 0.253 | 1.96 | 1.09 | 0.076 |
| fMDD [MDD] |  |  |  | -0.61 | 2.26 | 0.787 |
| **Random Effects** | | | | | | |
| σ^2^ | 221.73 | | | 214.74 | | |
| τ_00_ | 197.24 _ID_ | | | 128.38 _ID_ | | |
| τ_11_ | 260.05 _ID.fSnack1_ | | | 273.77 _ID.fSnack1_ | | |
|  | 581.17 _ID.fSnack2_ | | | 455.07 _ID.fSnack2_ | | |
|  | 832.24 _ID.fSnack3_ | | | 556.48 _ID.fSnack3_ | | |
|  | 733.98 _ID.fSnack4_ | | | 503.92 _ID.fSnack4_ | | |
|  | 198.60 _ID.fSnack5_ | | | 263.84 _ID.fSnack5_ | | |
|  | 1165.02 _ID.fSnack6_ | | | 646.69 _ID.fSnack6_ | | |
|  | 25.59 _ID.fPhase_dicho_FCR_TT1_ | | | 29.33 _ID.fPhase_dicho_FCR_TT1_ | | |
| ρ_01_ | 0.10 | | | 0.19 | | |
|  | -0.22 | | | -0.14 | | |
|  | 0.09 | | | 0.15 | | |
|  | -0.12 | | | -0.04 | | |
|  | 0.07 | | | 0.02 | | |
|  | 0.10 | | | -0.02 | | |
|  | 0.37 | | | -0.02 | | |
| ICC | 0.79 | | | 0.73 | | |
| N | 103 _ID_ | | | 103 _ID_ | | |
| Observations | 3605 | | | 3605 | | |
| Marginal R^2^ / Conditional R^2^ | 0.186 / 0.825 | | | 0.210 / 0.789 | | |
